# Genome-wide association study highlights 44 loci for transient ischemic attack and shared genetic architecture with stroke

**DOI:** 10.64898/2026.07.10.26357754

**Authors:** Shuyuan Hu, Ping Zhu, Shiyang Wu, Shan Gao, Ruibai Wang, Fengzhen Liu, Yijie He, Zhifa Han, Tao Wang, Mingxin Wang, Changhong Ren, Xunming Ji, Wenbo Zhao, Sijie Li, Guiyou Liu

## Abstract

Transient ischemic attack (TIA) is a critical harbinger of subsequent stroke, and most genetic risk remains uncharacterized. Here we firstly performed the largest TIA genome-wide association study (GWAS) meta analysis in 1,332,453 European individuals (58,976 cases and 1,273,477 controls), followed by an independent replication in 610,409 non-European individuals (23,557 TIA and 586,852 controls), a multi-ancestry GWAS meta analysis in 1,942,862 individuals (82,533 cases and 1,860,329 controls), and a cross-trait GWAS meta-analysis of TIA with stroke and its subtypes. We identified 44 loci including 25 known stroke loci and 19 TIA specific loci (*CELSR2*, *SLC4A7*, *CASC15*, *SRRM3*, *SLC44A1*, *LOC107984361*, *GSE1*, *LOC105372530*, *HCG20*, *OXR1*, *SLC4A1*, *RBBP8*, *TUSC3*, *DCC*, *PALMD*, *ZNF475*, *CTAGE1*, *FUT2* and *MRPS6*). Post-GWAS pinpointed 51 high confidence genes (24 are potential therapeutic targets) and 13 statistically significant pathways including protein-lipid complex, neurofibrillary tangle, high-density lipoprotein particle. These findings provide critical insights into the genetic basis of TIA.

## Introduction

Transient ischemic attack (TIA) is often referred to as a “mini stroke”. Unlike ischemic stroke that is defined as a sudden focal neurologic dysfunction with imaging-confirmed infarct, TIA is defined as a transient focal neurologic dysfunction caused by cerebral ischemia without imaging-confirmed infarct ^1,2^. TIA is a critical harbinger of subsequent stroke, with up to 17.8% of TIA patients progressing to stroke within 90 days, nearly half within the first 48 hours ^3^. Patients with TIA had a stroke cumulative incidence of 12.5% and 19.8% during the 5-year and 10-year follow-up periods, respectively ^4^. Meanwhile, TIA associated with persistent cognitive deficits, coronary artery disease, epilepsy, and disability/mortality risks, which underscored its profound clinical significance ^2,5–9^.

Genetic factors play important roles in the etiology of both TIA and ischemic stroke ^10^. It is estimated that the heritability of ischemic stroke is up to 38% ^11^. Until now, large-scale genome-wide association study (GWAS) datasets have identified common ischemic stroke genetic variants and more than 90 risk loci across diverse populations and subtypes ^12–16^. Unlike ischemic stroke, the majority of genetic risk of TIA remains uncharacterized until now. To address this gap, we firstly performed the largest TIA GWAS meta analysis in 3 cohorts including 1,332,453 individuals of European ancestry (58,976 cases and 1,273,477 controls) (stage 1), followed by an independent replication in 610,409 individuals of non-European ancestry (23,557 TIA and 586,852 controls) (stage 2), a multi-ancestry GWAS meta analysis in 1,942,862 individuals (82,533 cases and 1,860,329 controls) (stage 3), and a cross-trait GWAS meta-analysis of TIA and stroke as well as its subtypes (stage 4). We then characterized the genetic findings using kinds of post-GWAS approaches including gene mapping, gene-based association test, gene set enrichment analysis, multi-omics integrative analysis, genetic correlation analysis, phenome-wide association analysis, drug-gene interaction analysis, differential gene expression analysis, tissue and cell analysis. The overall study design is presented in Fig. 1.

**Fig. 1.**
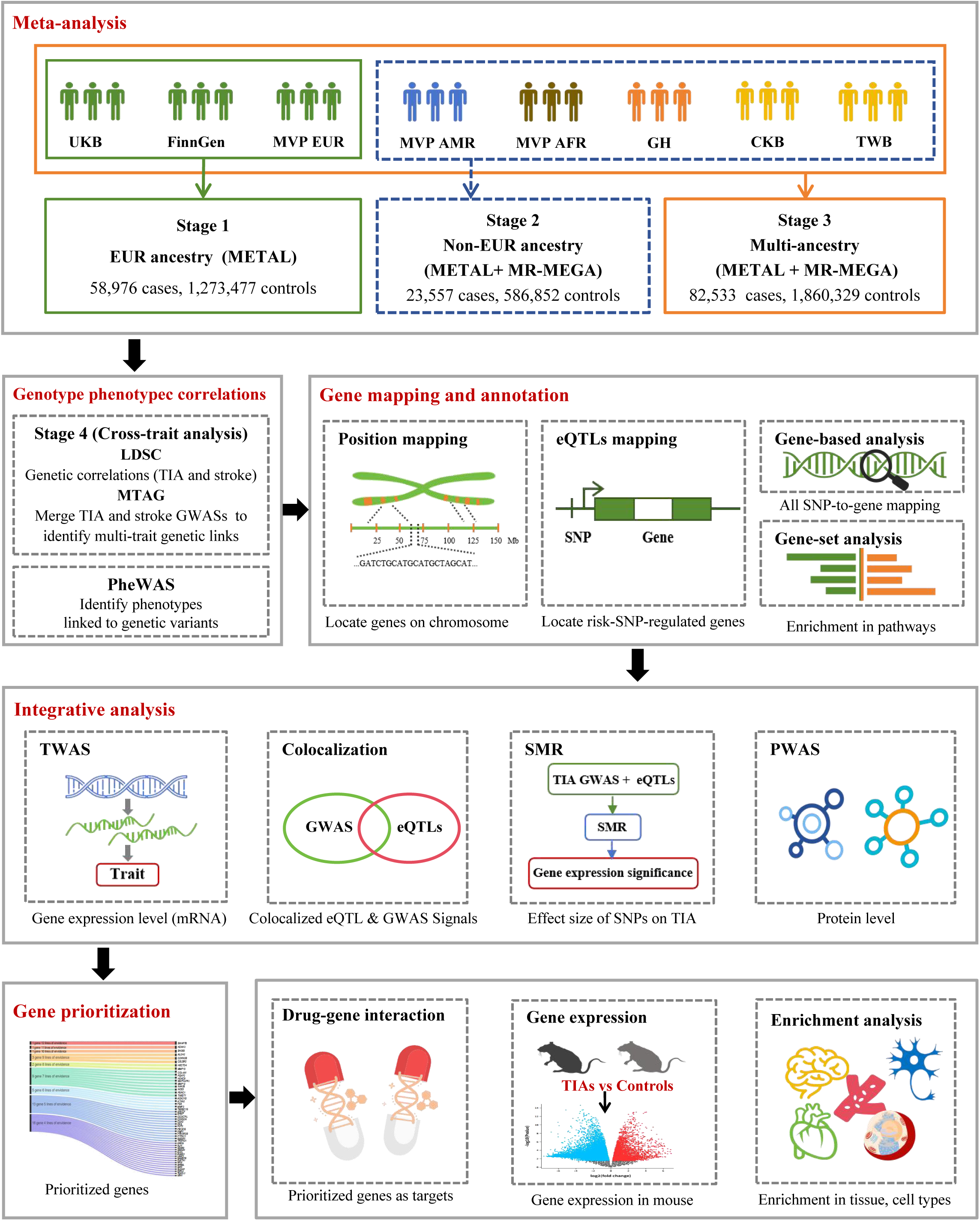
Overview of the study workflow. We performed the largest TIA GWAS meta analysis in 3 cohorts including 1,332,453 individuals of European ancestry (58,976 cases and 1,273,477 controls) (stage 1), followed by an independent replication in 5 cohorts including 610,409 individuals of non-European ancestry (23,557 TIA and 586,852 controls) (stage 2), a multi-ancestry GWAS meta analysis in 8 cohorts including 1,942,862 individuals (82,533 cases and 1,860,329 controls) (stage 3), and a cross-trait GWAS meta-analysis of TIA and stroke as well as its subtypes (MTAG, stage 4). We then characterized the genetic findings using kinds of post-GWAS approaches. LDSC was used to estimate the genetic correlation between TIA and stroke sand PheWAS to pinpoint the clinical phenotypes associated with the GWAS significant TIA genetic variants. Gene mapping (position mapping, eQTLs mapping) and gene-based association test were conducted to identify TIA risk genes. Several multi-omics integrative analyses, including TWAS, colocalization, SMR, and PWAS, were carried out to prioritize the TIA risk genes. Drug-gene interaction analysis was used to identify the potential therapeutic targets. Finally, LDSC-SEG and MAGMA were used to explore whether TIA heritability is enriched in specific tissues, cell types, or biological pathways. UKB, UK Biobank; EUR, European; MVP, Million Veteran Program; AMR, Hispanic/Latino American; AFR, African American; GH, Genes and Health; CKB, China Kadoorie Biobank; TWB, Taiwan Biobank; GWAS, genome-wide association study; MR-MEGA, Meta-Regression of Multi-AncEstry Genetic Association; LDSC, linkage disequilibrium score regression; MTAG, multitrait analysis of GWAS; PheWAS, phenome-wide association study; SNP, single nucleotide polymorphism; eQTLs, expression quantitative trait loci; TWAS, transcriptome-wide association study; SMR, summary-data-based mendelian randomization; PWAS, proteome-wide association study; LDSC-SEG, linkage disequilibrium score regression for specific gene expression; MAGMA, Multi-marker Analysis of GenoMic Annotation.

## Results

### European-specific GWAS meta-analysis (stage 1)

We conducted a TIA GWAS meta-analysis using a total of 58,976 TIA and 1,273,477 controls of European ancestry by combining 3 independent TIA GWAS datasets in participants of European ancestry from UK Biobank (UKB, 3,150 TIA and 417,381 controls, ICD-10 code G45) ^17^, FinnGen (R12 release, 24,948 TIA and 453,276 controls from Finnish, ICD-10 code G45) ^18^, and Million Veteran Program (MVP) European participants (30,878 TIA and 402,820 controls, Phecode 433.31) ^19^ (Supplementary Table 1a).

Using linkage disequilibrium score regression (LDSC) ^20^, we found statistically significant positive correlations across these three TIA GWAS datasets (Supplementary Table 2), which suggested that it was appropriate and reliable to combine these datasets in the meta-analysis. Using the fixed-effects inverse variance weighted (IVW) method implemented in METAL ^21^, the linkage disequilibrium (LD) information from the 1000 Genomes Project Phase 3 European ancestry reference panel implemented in Functional Mapping and Annotation (FUMA) with the default settings, we revealed 10 independent genome-wide significant risk loci including *CDKN2B-AS1* (rs7859727), *SH2B3* (rs7310615), *MMP3* (rs662558), *NOS3* (rs3918226), *HDAC9* (rs2023936), *PIK3CG* (rs17398575), *SWAP70* (rs415895), *FOXF2* (rs1922928), *CELSR2* (rs660240) and *KCNK3* (rs736699) (Fig. 2a, Table 1, Supplementary Table 3a). These 10 single nucleotide polymorphisms (SNPs) corresponding to these 10 loci are all common variants with the minor allele frequency (MAF) ≥1%. In particular, rs7859727 around the *CDKN2B-AS1* exhibited the most significant association with TIA (*P*=1.44E-18), and the remaining risk variants also demonstrated genome-wide significance (Fig. 3). Independent significant SNPs (*r*^2^<0.6) and lead SNPs (*r*^2^<0.1) around these 10 loci were provided in Supplementary Tables 4-5 using the LD information from the 1000 Genomes Project phase 3 European ancestry reference panel. The genome-wide SNP-based heritability (*h*²) on the liability scale was estimated at 0.034 (s.e.=0.002) assuming the TIA population prevalence 2.3% ^22^. Although we observed evidence of genetic inflation with a genomic inflation factor λ_GC_=1.263 (quantile-quantile (QQ) plot, Fig. 4a), LDSC intercept 1.054 (s.e.=0.008) indicated that the observed inflation was due to polygenicity rather than bias from population stratification.

**Fig. 2.**
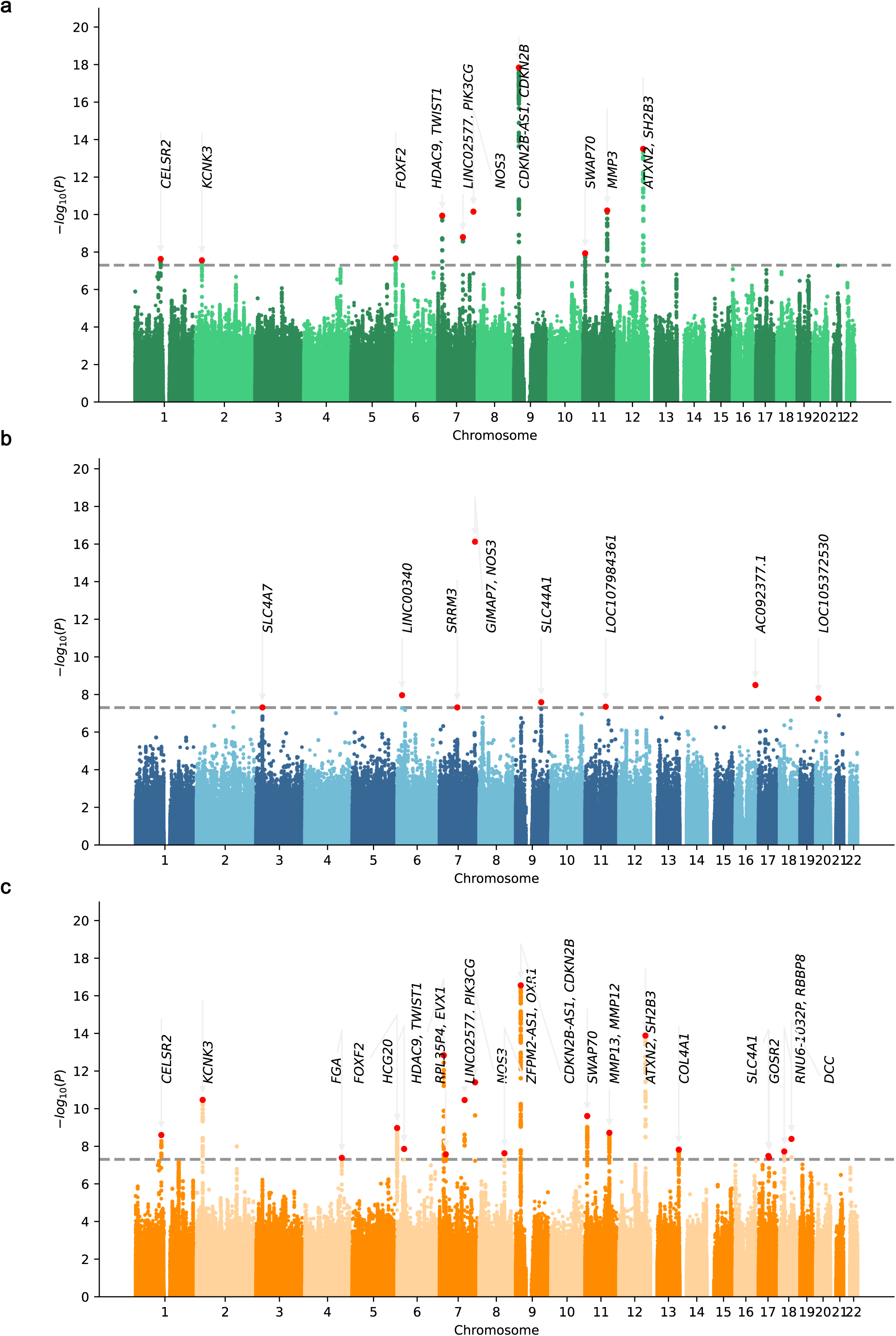
Manhattan plots of 3-stage TIA GWAS meta-analyses using METAL. **a.** Manhattan plot of stage 1 European-specific GWAS meta-analysis using 58,976 TIA and 1,273,477 controls. rs7859727 near *CDKN2B-AS1* showed the most significant association. **b.** Manhattan plot of stage 2 Non-European GWAS meta-analysis using 23,557 TIA and 586,852 controls. rs73159770 near *NOS3* showed the most significant association. **c.** Manhattan plot of stage 3 multi-ancestry (European and Non-European) GWAS meta-analysis using 82,533 TIA and 1,860,329 controls. rs7857118 near *CDKN2B-AS1* showed the most significant association. The *x* axis depicts the chromosome and base pair positions of genetic variants. The *y* axis shows the -log_10_(*P* values). *P* values are from the fixed-effects inverse variance weighted method implemented by METAL. All statistical tests were two-sided. The grey dash line shows the genome-wide significance threshold (*P*<5.00E-08).

**Table 1.**
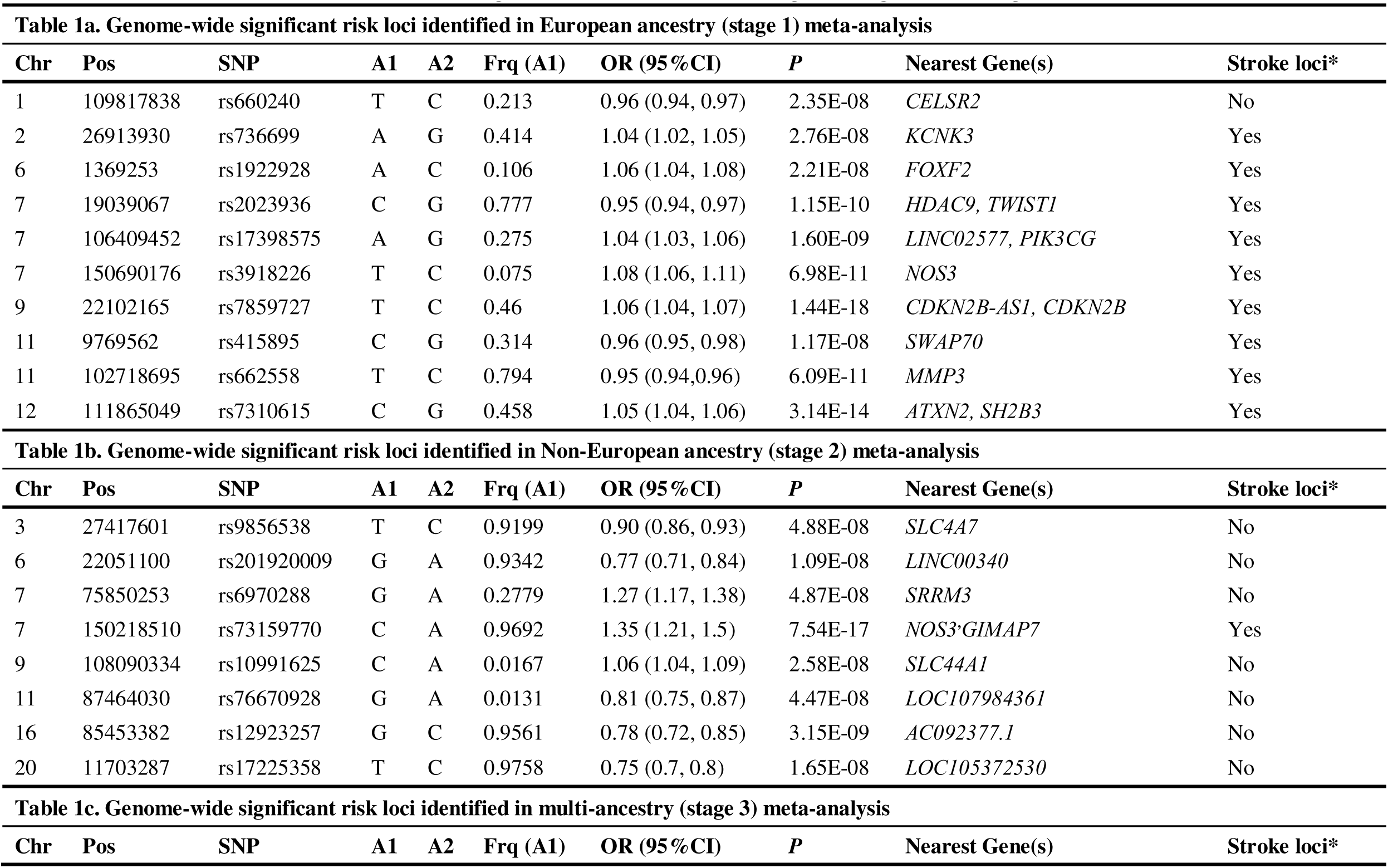
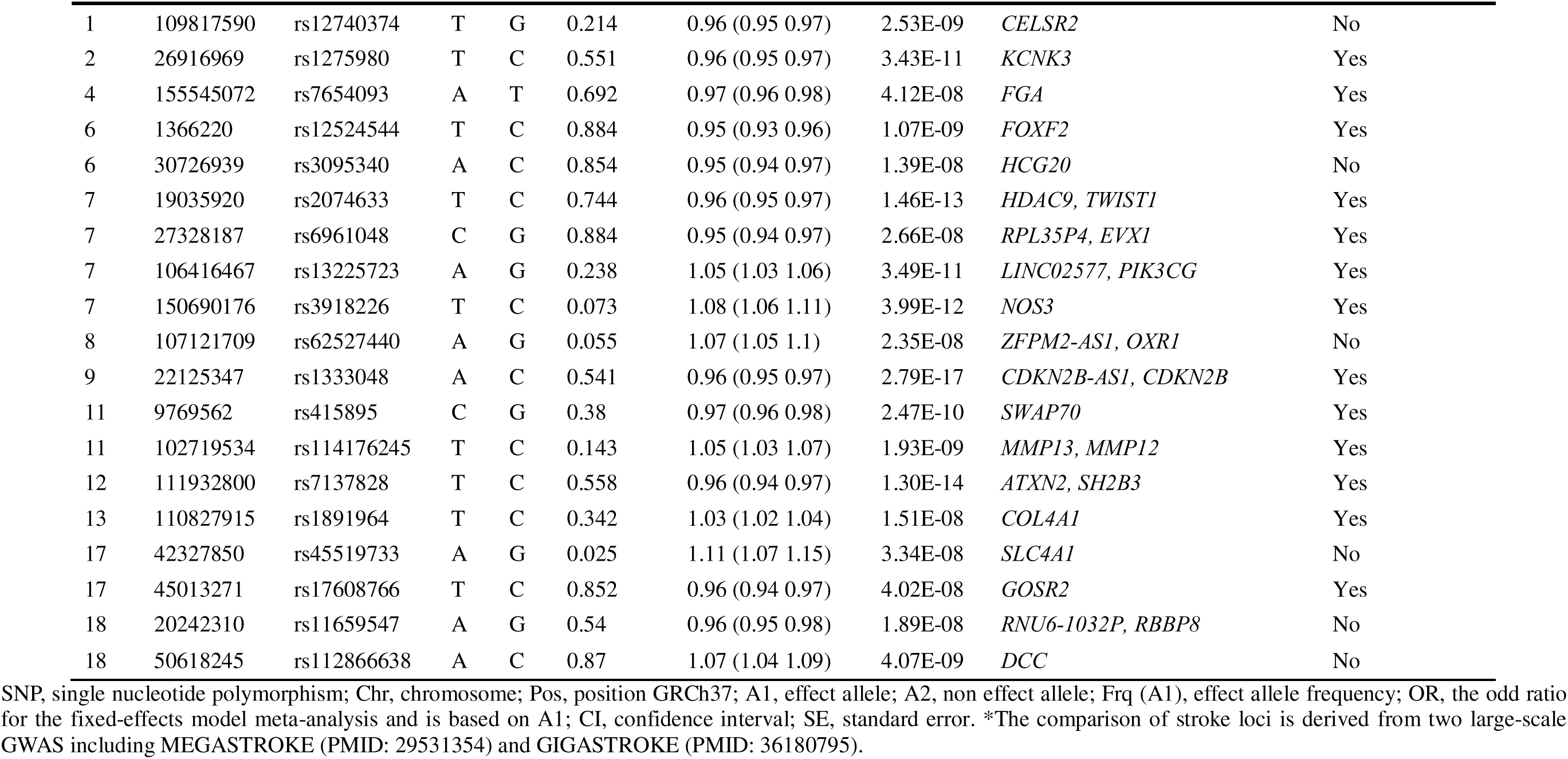
Genome-wide significant TIA loci from stage 1, stage 2 and stage 3 (METAL)

**Fig. 3.**
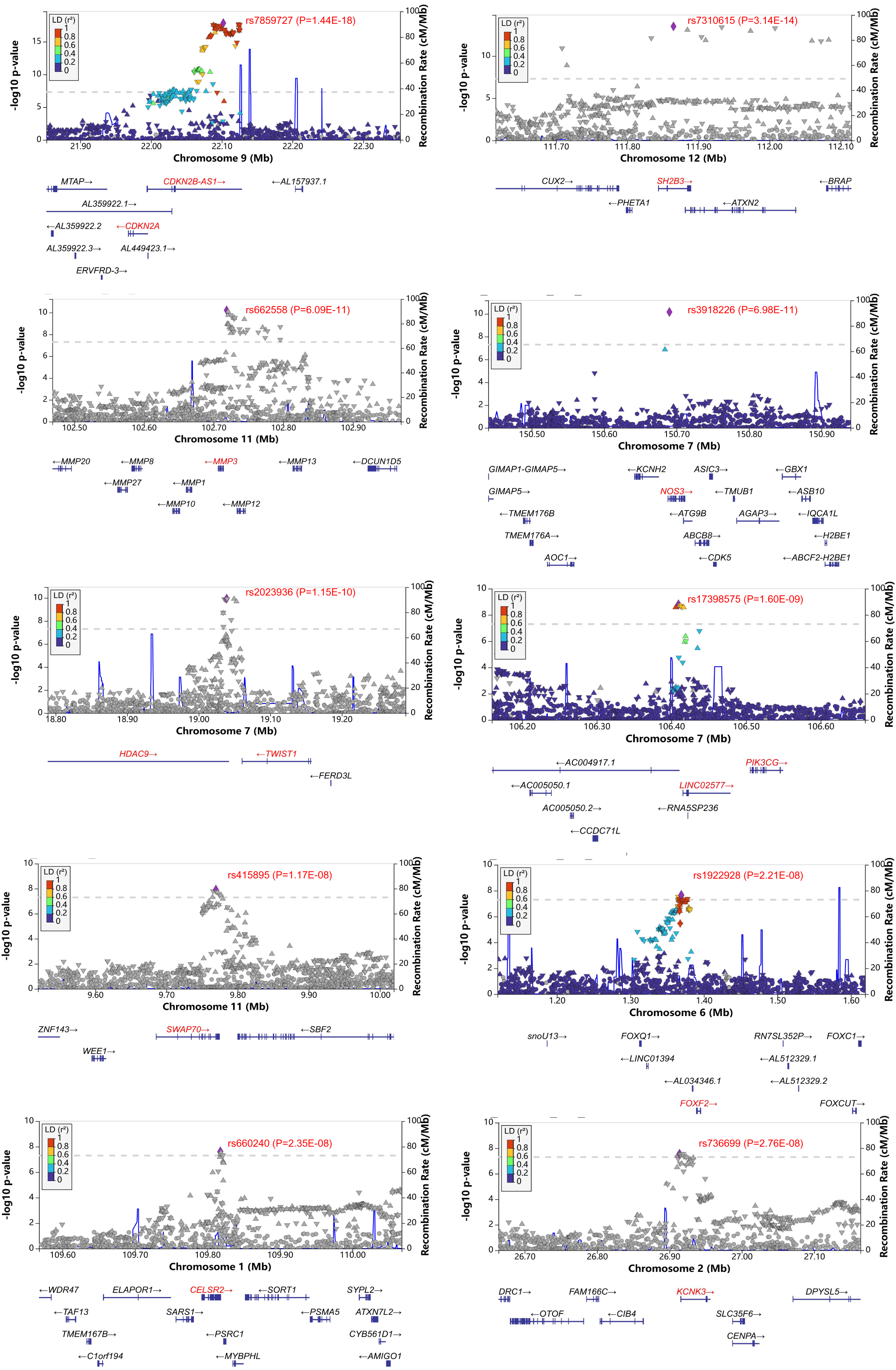
Locuszoom plots of 10 genome-wide significant TIA loci from European-specific meta-analysis (stage 1). LocusZoom plots of rs7859727 (*CDKN2B-AS1*), rs7310615 (*SH2B3*), rs662558 (*MMP3*), rs3918226 (*NOS3*), rs2023936 (*HDAC9*), rs17398575 (*LINC02577*), rs415895 (*SWAP70*), rs1922928 (*FOXF2*), rs660240 (*CELSR2*) and rs736699 (*KCNK3*). The *x* axis depicts the chromosome and base pair positions of genetic variants. The *y* axis shows the -log_10_(*P* values). *P* values are from the fixed-effects inverse variance weighted method implemented by METAL (v 2011-03-25). All statistical tests were two-sided. The grey dash line shows the genome-wide significance threshold (*P*<5.00E-08).

**Fig. 4.**
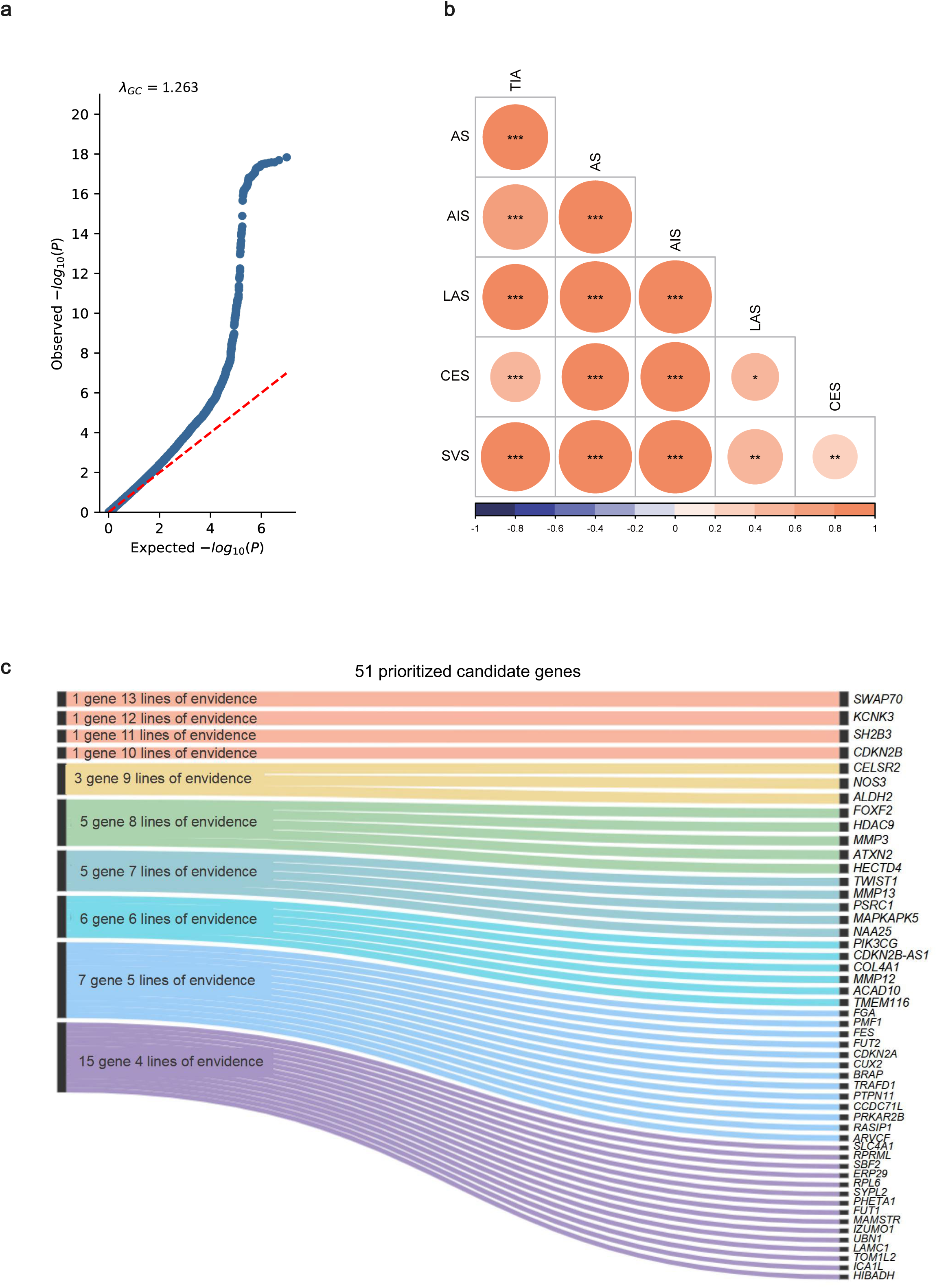
QQ Plot, LDSC and gene prioritization results. **a.** QQ plot and Lambda GC of the stage 1 European-specific meta-analysis. The observed *P* values were from the fixed-effects inverse variance weighted method implemented by METAL, and all statistical tests were two-sided. The red dashed line represents the predicted *P* value, and the blue framed line shows the 95% confidence interval. **b.** Genetic correlations between TIA from the stage 1 European-specific meta-analysis with stroke phenotypes from MEGASTROKE consortium (European ancestry). The genetic correlation coefficient was calculated using LDSC and is denoted by color scale from -1 (modena; negatively (anti-) correlated) to +1 (orange; positively correlated). * two-sided *P*L<L0.05, ** two-sided *P*L<L0.01 and *** two-sided *P*L<L0.001. **c.** Gene prioritization results. A total of 17 lines of evidence were used, and each column represents a type of supportive evidence. Genes supported by at least four lines of evidence are considered as high-confidence candidate genes. 51 genes were supported by at least 4 lines of evidence.

We further mapped the independent significant SNPs and SNPs in LD (*r*^2^≥0.6) (Supplementary Table 6) to their corresponding genes using both positional mapping (within 10 kilobases (kb)) and expression quantitative trait loci (eQTL) mapping (within 1 megabases (Mb)). Using ANNOVAR implemented in FUMA with the default settings for positional mapping ^23^, we identified 30 genes (Supplementary Table 7). eQTL mapping using the Genotype-Tissue Expression (GTEx) (version 10, primarily in individuals of European ancestry) ^24^ and MetaBrain (2,683 individuals of European ancestry) datasets ^25^, we identified 173 genes in human brain, blood vessels, and whole blood tissues with significant *P*<0.05 (Supplementary Table 8). In total, 27 genes were identified by both mapping methods, of which 9 genes including *CELSR2*, *KCNK3*, *FOXF2*, *HDAC9*, *TWIST1*, *NOS3*, *SWAP70*, *MMP3*, and *SH2B3* were located within the genome-wide significant loci.

### Independent replication in non-European (stage 2)

We replicated the stage 1 findings by performing a TIA GWAS meta-analysis in 610,409 non-European individuals (23,557 TIA and 586,852 controls) using five independent cohorts from MVP Hispanic/Latino American (MVP AMR, 2,792 TIA and 55,391 controls, phecode 433.31) ^19^, MVP African American (MVP AFR, 7,606 TIA and 109,704 controls, phecode 433.31) ^19^, China Kadoorie Biobank (CKB, East Asian, 2,742 TIA and 73,812 controls, ICD-10 code G45) ^26^, Taiwan Biobank (TWB, East Asian, 9,928 TIA and 304,660 controls, ICD-10 code G45, and phecode 433.31) ^27^, and Genes and Health (GH, South Asian, 489 TIA and 43,285 controls, ICD-10 code G45) (Supplementary Table 1b) ^28^.

Using the fixed-effects IVW implemented in METAL ^21^ and the LD information from the 1000 Genomes multi-ancestry reference panel implemented in FUMA with the default settings, we identified 8 independent genome-wide significant loci by confirming *NOS3* (rs73159770) from stage 1 and highlighting 7 additional loci which indicated the ethnic differences in TIA heritability (*P*<5E-08, Supplementary Table 3b, Extended Fig.1). 7 of 10 loci in stage 1 were replicated in stage 2 (*P*<0.05). Briefly, 7 of the 10 lead variants corresponding to 10 loci were nominally associated with TIA (*P*<0.05) in at least one single replication dataset and/or meta-analysis dataset, with the same direction in at least two replication datasets. Notably, rs736699 (*KCNK3*), rs2023936 (*HDAC9*), and rs415895 (*SWAP70*) showed consistent directions in 5, 4 and 5 replication datasets, respectively (Supplementary Table 9a). Given the different ancestral backgrounds of the contributing cohorts, a sensitivity meta-analysis was performed using a multi-ancestry meta-analysis approach implemented in MR-MEGA (Meta-Regression of Multi-AncEstry Genetic Association) ^29^. MR-MEGA did not identify additional genome-wide significant loci, but further confirmed rs736699 (*KCNK3*), rs2023936 (*HDAC9*), and rs415895 (*SWAP70*) (suggestive *P*<0.05, Supplementary Table 9b).

Independent significant SNPs (*r*²<0.6) and lead SNPs (*r*²<0.1) were defined using LD data from the 1000 Genomes Project Phase 3 multi-ancestry panel (Supplementary Tables 10-11). Using these independent significant SNPs and SNPs in LD (Supplementary Table 12), we identified 7 genes using positional mapping (Supplementary Table 13) and 8 genes using eQTL mapping in brain, blood vessels, and whole-blood from GTEx v10 (Supplementary Table 14).

### Multi-ancestry GWAS meta-analysis (stage 3)

We further conducted a cross-ancestry meta-analysis by combining 8 independent GWAS datasets from both stage 1 (UKB, FinnGen and MVP European) and stage 2 (MVP AMR, MVP AFR, CKB, TWB, and GH) including 1,942,862 individuals (82,533 TIA and 1,860,329 controls) (Supplementary Table 1c). Using the fixed-effects IVW implemented in METAL ^21^ and the LD information from the 1000 Genomes multi-ancestry panel implemented in FUMA with default settings, we confirmed all 10 genome-wide significant loci from stage 1 European-specific GWAS meta-analysis including *CDKN2B-AS1*, *SH2B3*, *MMP13, HDAC9*, *NOS3*, *PIK3CG*, *SWAP70*, *FOXF2, CELSR2* and *KCNK3*. Meanwhile, we identified 9 additional novel loci including *DCC*, *HCG20*, *COL4A1*, *RBBP8*, *ZFPM2-AS1*, *EVX1*, *SLC4A1*, *GOSR2* and *FGA* (Fig 2b, Table 1, Supplementary Figs. 1-19). Four loci showed homogeneous effects across all these 8 datasets including *SWAP70*, *FGA*, *COL4A1* and *EVX1*. *CDKN2B-AS1* was still the most significant signal tagged by rs7857118 (*P*[=2.79E-17) (Table 1, Supplementary Table 3c). Supplementary Tables 15-16 provided the independent significant SNPs (*r*^2^<0.6) and lead SNPs (*r*^2^<0.1) around these loci using the LD information from the 1000 Genomes multi-ancestry reference panel, respectively. We observed evidence of genetic inflation with λ_GC_=1.233 (QQ plot, Supplementary Fig. 20). LDSC intercept 1.037 (s.e.=0.007) further supported that the observed inflation was due to polygenicity rather than bias from population stratification.

MR-MEGA identified 12 independent genome-wide significant TIA loci by confirming 10 of 19 loci from METAL and revealing 2 additional novel loci (*TUSC3* and *LDLR*; Fig. 5a, Supplementary Figs. 21-22, Supplementary Tables 17-18). Of the 10 loci, 8 (*KCNK3*, *FOXF2*, *HDAC9*, *PIK3CG*, *NOS3*, *SWAP70*, *MMP13*, and *SH2B3*) showed consistent effects across ancestries, while 2 (*CDKN2B-AS1* and *SLC4A1*) exhibited ancestry-correlated heterogeneity. Crucially, no variants showed significant residual heterogeneity, indicating that the associations are not substantially confounded by differences in study design, phenotyping, or exposures (Supplementary Tables 17-18). 2 novel loci exhibited significant ancestral heterogeneity (*P* _value_ancestry_het_<0.05), but no residual heterogeneity (*P*_value_residual_het_ >0.05). MR-MEGA did not provide the effect sizes other than for each ancestry component, and the subsequent downstream analyses were conducted using GWAS results from METAL.

**Fig. 5.**
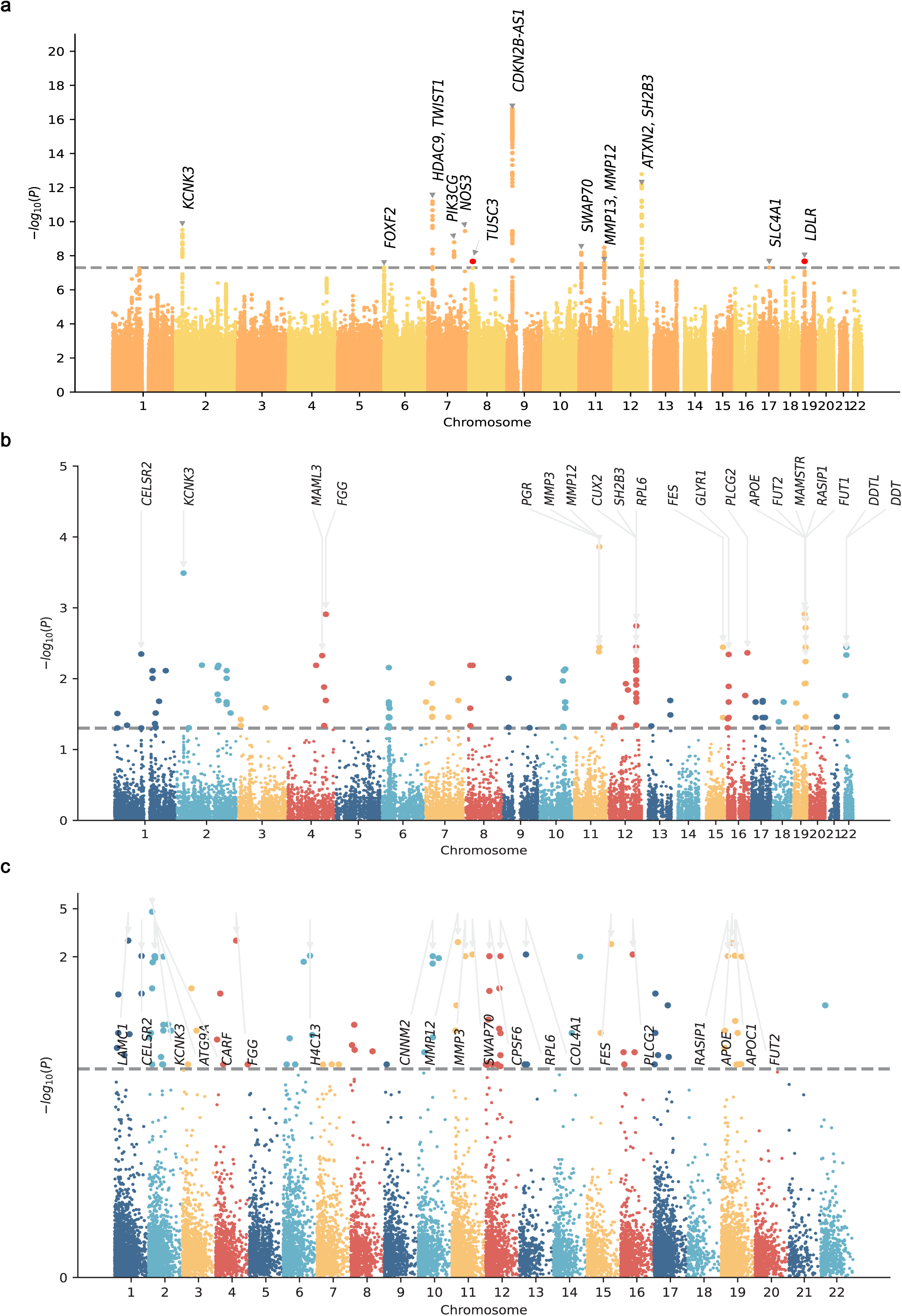
Manhattan plots of cross-ancestry MR-MEGA meta-analysis and gene-based analysis for TIA. **a.** Manhattan plot of TIA MR-MEGA (stage 3). The *x* axis represents the chromosomal positions and the *y* axis shows the -log_10_(*P* values), and all statistical tests were two-sided. The horizontal dashed line indicates the genome-wide significance threshold (*P*<5.00E-08), and red-colored markers denote two additional loci identified by MR-MEGA. **b.** Gene-based analysis of TIA European ancestry (stage 1). The *y* axis shows -log_10_*P* of F statistics (twosided nominal *P* values) implemented in MAGMA. The grey line in dicates false discovery rate (FDR) -corrected genome-wide significance *P* threshold (*P*<0.05). The Figure presents the top 20 genes. **c.** Gene-based analysis of TIA multi-ancestry (stage 3, METAL). The *y* axis shows -log_10_*P* of F statistics (two sided nominal *P* values) implemented in MAGMA. The grey line indicates FDR-corrected genome-wide significance *P* threshold (*P*<0.05). The Figure presents the top 20 genes.

We identified 42 genes using positional mapping (Supplementary Table 19) and 310 genes using eQTL mapping in brain, blood vessels, and whole-blood from GTEx v10 (primarily in individuals of European ancestry) and MetaBrain (2,683 individuals of European ancestry) (Supplementary Table 20). Twenty-nine genes were detected by both approaches, 14 of which were located within the genome-wide significant loci including *CELSR2*, *KCNK3*, *FOXF2*, *HDAC9*, *TWIST1*, *NOS3*, *CDKN2B*, *SWAP70*, *MMP13*, *SH2B3*, *COL4A1*, *SLC4A1*, *GOSR2*, and *DCC*.

### Cross-trait GWAS meta-analysis of TIA and stroke (stage 4)

We first investigated the genetic correlations of TIA (stage 1 European) with stroke, ischemic stroke and its three subtypes including large-artery atherosclerotic stroke (LAS), cardioembolic stroke (CES), and small-vessel stroke (SVS) from the MEGASTROKE consortium (European ancestry, no sample overlap with TIA) ^30^ using LDSC (v1.0.1) with the default settings, and LD information from the 1000 Genomes Project phase 3 European reference panel ^23^. We found that TIA showed statistically significant positive genetic correlations with stroke and its subtypes, especially ischemic stroke demonstrating the strongest positive genetic correlation (*rg*=0.96, SE=0.03, *P*<8.69E-308) (Fig. 4b and Supplementary Table 21).

We further conducted a cross-trait GWAS meta-analysis of TIA (stage 1 European) and stroke as well as its subtypes (ischemic stroke, LAS, CES and SVS in European) from the MEGASTROKE consortium ^30^ using the multitrait analysis of GWAS (MTAG), which boosts the effective sample size and statistical power to detect genetic associations for each trait ^31^. MTAG significantly increased the effective sample sizes of the TIA GWAS (from 1,332,453 to 1,742,764, 30.8%) and stroke GWAS as provided in Supplementary Table 22. MTAG discovered 28 independent genome-wide significant TIA loci by confirming 12 loci from single trait TIA GWAS, and highlighting 16 novel loci (Fig. 6a-e, Supplementary Table 23). MTAG accordingly enhanced stroke genetics by identifying 36 loci associated with stroke or its subtypes, of which 8 are novel loci (Fig. 6f-j, Supplementary Table 24). The top and third significant loci in GIGASTROKE ^13^, *PITX2* and *ZFHX3,* were genome-wide significant loci associated with TIA and stroke using MTAG, respectively (Supplementary Table 24). In total, our four-stage GWAS uncovered 44 unique genome-wide significant TIA loci with 10 identified in stage 1, 8 in stage 2, 21 in stage 3, and 28 in stage 4 (Supplementary Table 25).

**Fig. 6.**
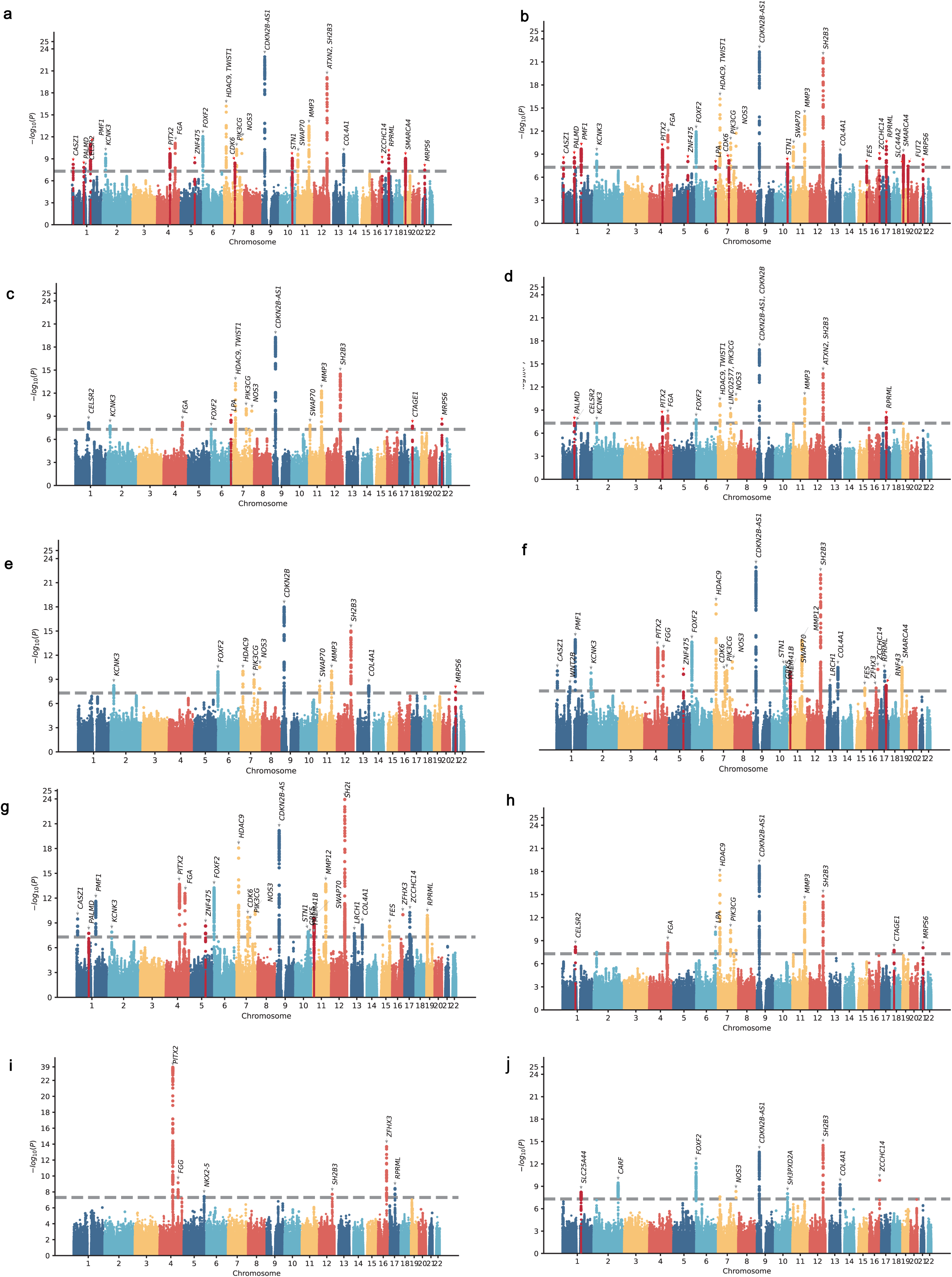
Manhattan plots of GWAS results using MTAG. **a.** Manhattan plots of TIA GWAS results by integration with stroke GWAS using MTAG. **b.** Manhattan plots of TIA GWAS results by integration with ischemic stroke GWAS using MTAG. **c.** Manhattan plots of TIA GWAS results by integration with LAS GWAS using MTAG. **d.** Manhattan plots of TIA GWAS results by integration with CES GWAS using MTAG. **e.** Manhattan plots of TIA GWAS results by integration with SVS GWAS using MTAG. **f.** Manhattan plots of stroke GWAS results by integration with TIA GWAS using MTAG. **g.** Manhattan plots of ischemic stroke GWAS results by integration with TIA GWAS using MTAG. **h.** Manhattan plots of large-artery atherosclerotic stroke (LAS) GWAS results by integration with TIA GWAS using MTAG. i. Manhattan plots of cardioembolic stroke (CES) GWAS results by integration with TIA GWAS using MTAG. **j.** Manhattan plots of small-vessel stroke (SVS) GWAS results by integration with TIA GWAS using MTAG. The *x* axis represents chromosomal positions, the *y* axis shows -log10(*P*-values), and the horizontal dashed line indicates the genome-wide significance threshold (*P*<5E-08). Red markers indicate novel loci identified via MTAG that were not detected in the original single-trait GWAS.

### Genetic association of TIA loci with stroke

We conducted a comparative analysis of current TIA loci with known stroke loci by evaluating the genetic association of 44 TIA loci with stroke and its subtypes using large cross-ancestry GWAS datasets from the MEGASTROKE ^30^ and GIGASTROKE consortia ^13^. At the genetic variant level, 25 lead variants corresponding to 44 loci were associated with stroke and/or its subtypes (Bonferroni-corrected *P*<0.05/44=1.14E-03; Supplementary Table 26). At the locus level, 25 of the 44 TIA loci were known stroke loci (e.g., *KCNK3*, *FGA* and *CDKN2B-AS1*), while the remaining 19 were TIA-specific loci including *CELSR2*, *SLC4A7*, *CASC15*, *SRRM3*, *SLC44A1*, *LOC107984361*, *GSE1*, *LOC105372530*, *HCG20*, *OXR1*, *SLC4A1*, *RBBP8*, *TUSC3*, *DCC*, *PALMD*, *ZNF475*, *CTAGE1*, *FUT2* and *MRPS6* (Supplementary Table 25). These findings highlighted both shared and distinct genetic etiologies in TIA and stroke.

### Phenome-wide association study

In addition to stroke, we conducted a phenome-wide association study (PheWAS) of the lead variants corresponding to each TIA locus using a broad range of phenotypes including 1400 traits from UKB ^32^, 2,068 traits from MVP ^19^, and 2,469 traits from FinnGen R12 ^33^. These variants were mainly associated with coronary artery disease, coronary atherosclerosis, hypertension, systolic blood pressure, diastolic blood pressure, total cholesterol, high density lipoprotein cholesterol, and low density lipoprotein cholesterol (Supplementary Tables 27-58). Briefly, PheWAS confirmed known biology, which further supported our genetic findings.

### Gene-based association test and gene set enrichment analysis

Firstly, we conducted a gene-based association test of TIA GWAS meta-analysis summary data using MAGMA 34 and LD information from the 1000 Genomes Project phase 3 European ancestry reference panel. In stage 1, we identified 127 statistically significant genes after a false discovery rate (FDR) correction *P*<0.05 (Fig. 5b, Supplementary Table 59). Seventeen genes (*CELSR2*, *KCNK3*, *CDKN2B*, *MMP3*, *CUX2*, *SH2B3*, *ATXN2*, *BRAP*, *ACAD10*, *ALDH2*, *MAPKAPK5*, *TMEM116*, *ERP29*, *NAA25*, *TRAFD1*, *HECTD4* and *PTPN11*) were shared across the positional mapping, eQTL mapping and MAGMA. Specifically, *CELSR2*, *KCNK3*, *MMP3* and *SH2B3* were located within the genome-wide significant loci. In stage 2, no statistically significant genes (FDR<0.05) were detected. In stage 3, we identified 102 statistically significant genes (FDR<0.05) (Fig. 5c, Supplementary Table 60). Sixty-six genes were shared in stage 1 and stage 3, and six genes (*KCNK3*, *FGA*, *MMP12*, *COL4A1*, *CELSR2* and *SH2B3*) were located within genome-wide significant loci. Eleven genes (*KCNK3*, *SWAP70*, *MMP3*, *SH2B3*, *ACAD10*, *ALDH2*, *MAPKAPK5*, *NAA25*, *HECTD4*, *COL4A1* and *CELSR2*) were shared across the positional mapping, eQTL mapping and MAGMA. *CELSR2*, *KCNK3*, *SWAP70*, *SH2B3* and *COL4A1* were located within the genome-wide significant loci.

Secondly, we conducted a gene set enrichment analysis of TIA GWAS meta-analysis summary data using MAGMA. In stage 1, we identified 4 statistically significant gene ontology (GO) pathways (FDR<0.05) including protein-lipid complex (FDR=3.46E-03), negative regulation of multicellular organismal process (FDR=1.02E-02), neurofibrillary tangle (FDR=2.56E-02), and high-density lipoprotein particle (FDR=4.08E-02). In stage 2, we identified 2 statistically significant GO pathways (FDR<0.05) including protein-lipid complex (FDR=8.44E-03) and high-density lipoprotein particle (FDR=4.75E-02). In stage 3, we identified 12 statistically significant GO pathways (FDR<0.05) including negative regulation of multicellular organismal process (FDR=2.80E-02), negative regulation of alcohol biosynthetic process (FDR=2.80E-02), protein-lipid complex (FDR=2.76E-02), cardiac neural crest cell migration involved in outflow tract morphogenesis (FDR=2.80E-02), and high-density lipoprotein particle (FDR=3.32E-02). Here, we present the top 10 significant GO terms including biological process, molecular function, and cellular component in each stage in Supplementary Tables 61-63 and Extended Fig. 2a-c, respectively.

### Transcriptome-wide association study and colocalization analysis

We conducted a transcriptome-wide association study (TWAS) to identify risk genes whose expression levels were associated with TIA by integrating the stage 1 TIA GWAS meta-analysis summary data with eQTL datasets from GTEx v8 (13 brain tissues, 3 vessel tissues, 2 heart tissues, and whole blood) ^24^, and Common Mind Consortium (CMC) brain tissue ^35^. We pinpointed 190 significant genes (FDR<0.05), with *CDKN2A* showing the strongest association in brain cortex (*z*=6.19, *P*=5.88E-10). Genome-wide significant loci *KCNK3*, *SWAP70* and *CDKN2B-AS1* exhibited significant associations across multiple tissues, including *KCNK3* in heart atrial appendage (*z*=-5.20, *P*=1.95E-07), brain caudate (*z*=-4.09, *P*=4.34E-05), brain putamen (*z*=-4.21, *P*=2.55E-05), and brain cortex (*z*=-3.64, *P*=2.77E-04), *SWAP70* in tibial artery (*z*=4.52, *P*=6.04E-06), and *CDKN2B-AS1* in CMC brain (*z*=5.43, *P*=5.75E-08) (Fig. 7a-b, Supplementary Table 64).

**Fig. 7.**
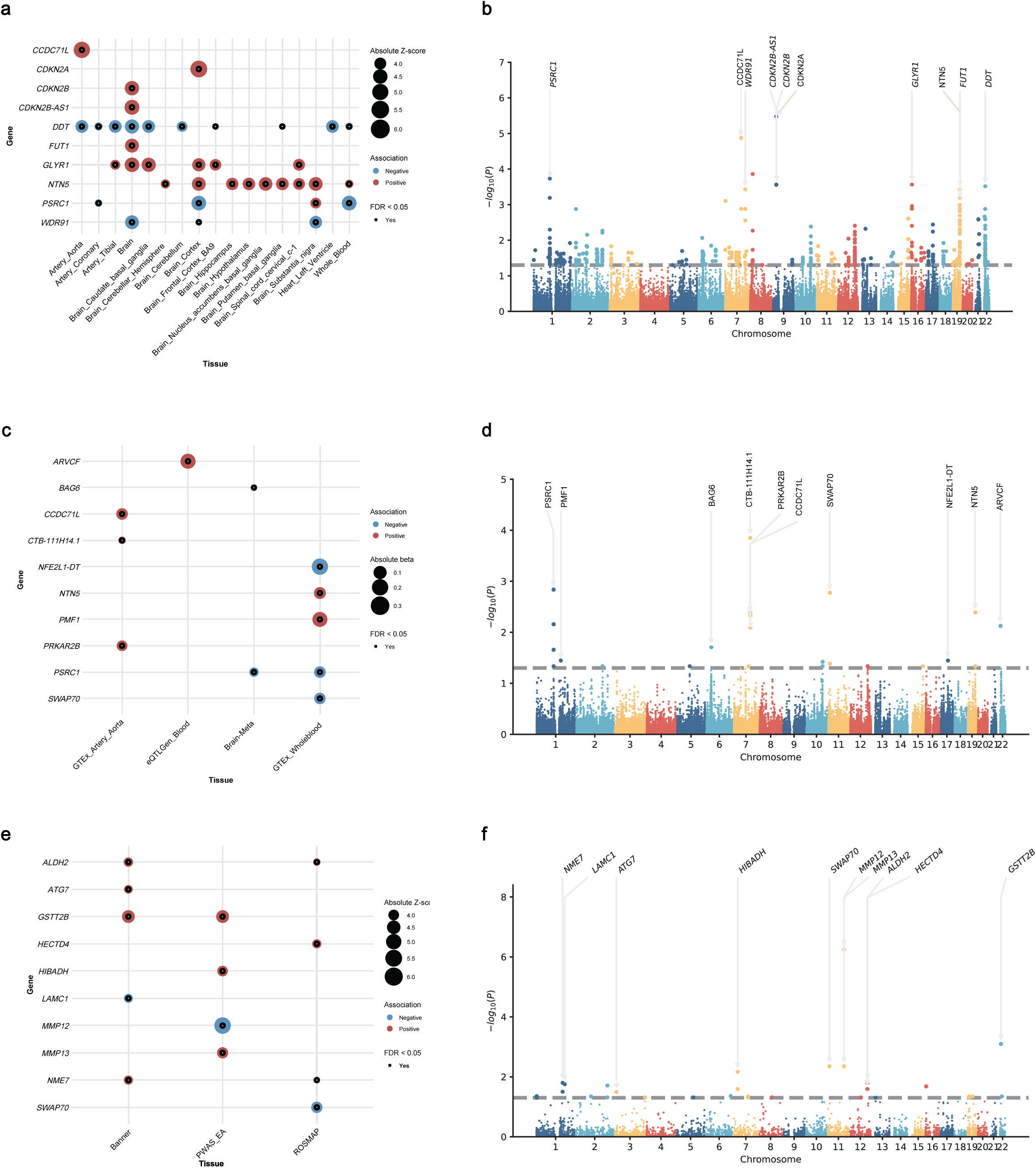
Bubble plots and Manhattan plots of TWAS, SMR, and PWAS results. **a.** Bubble plots of TWAS, which displays gene-tissue associations. *X* axis, tissues; *y* axis, candidate genes. Bubble color reflects the association direction (red=positive, blue=negative), size corresponds to absolute Z-score, and black dots mark FDR<0.05. **b.** Manhattan plots of TWAS. *X* axis, chromosomal position; *y* axis, −log_10_(*P* values). **c.** Bubble plots of SMR. *X* axis, tissues; *y* axis, candidate genes. Bubble attributes (color, size, significance dots) follow the same logic as panel Fig.7a. **d.** Manhattan plots of SMR. *X* axis, chromosomal position; *y* axis, −log_10_(*P* values). **e**. Bubble plots of PWAS. *X* axis, tissues; *y* axis, candidate genes. Bubble features align with panels Fig.7a/c. **f.** Manhattan plots of PWAS. *X* axis, chromosomal position; *y* axis, −log_10_(*P* values). All Manhattan plots shows -log_10_(FDR) on the *y* axis, with the grey dashed line at -log_10_(0.05) marking the FDR significance threshold (FDR<0.05). All plots presents the top 10 genes. All statistical tests were two-sided.

To determine whether the TIA GWAS signals and eQTL signals were influenced by the same genetic variants, we performed a colocalization analysis using COLOC ^36,37^ implemented in FUSION ^38^. Colocalization analysis using GTEx v8 ^24^, CMC ^35^ eQTLs and TIA GWAS datasets showed the shared causal variants for 31 genes (with a posterior probability of PPH4[≥[0.8) ^39–41^, including *KCNK3*, *ALDH2* and *SWAP70*. *SWAP70* remained the top-ranked gene, demonstrating colocalization specifically in tibial artery (PP4=0.999). Colocalization was observed for *CCDC71L*, *PRKAR2B* and *ARVCF* in aorta artery (PP4=0.998, 0.998, and 0.993, respectively), *PSRC1* in brain cortex (PP4=0.990) and whole blood (PP4=0.989) (Supplementary Table 65).

### Summary-data-based Mendelian randomization

TWAS could not determine whether TWAS signals have the causal effects on TIA ^42^. Here, we selected summary-data-based mendelian randomization (SMR) ^43^ to further investigate the causal effects of gene expression on TIA by integrating the stage 1 TIA GWAS meta-analysis summary data with eQTL data from GTEx v8 (13 brain tissues, 3 vessel tissues, 2 heart tissues, and whole blood) ^24^, eQTLGen (whole blood) ^44^ and BrainMeta v1 (brain tissue) ^45^. We identified 20 statistically significant genes with FDR-corrected *P*<0.05, and *P* _Heterogeneity in the dependent instrument (HEIDI) test_ >0.01. Fifteen genes were shared in both TWAS and SMR including *CCDC71L*, *PSRC1*, *NTN5*, *RASIP1*, *PRKAR2B*, *ARVCF*, *FES*, *AMIGO1*, *SWAP70*, *MSH3*, *PPP1R9A*, *COL17A1*, *MAPKAPK5-AS1*, *PMF1* and *SH3PXD2A*. Specifically, *PSRC1* was idenfieid in both brain and blood, *SWAP70* and *SH2B3* exclusively in blood (Fig. 7c-d and Supplementary Table 66).

### Proteome-wide association study

We conducted a proteome-wide association study (PWAS) to integrate the stage 1 TIA GWAS meta-analysis summary data with three human brain and blood protein QTL (pQTL) datasets: Banner Sun Health Research Institute (Banner) ^46^, Religious Orders Study and Rush Memory and Aging Project (ROS/MAP) ^47^, and Atherosclerosis Risk in Communities study (ARIC) ^48^. We identified 25 statistically significant genes with FDR<0.05 including *GSTT2B* (Banner, ARIC), *ALDH2* (Banner, ROSMAP), *NME7* (Banner, ROSMAP), *PANK4* (Banner, ROSMAP) and *PON2* (Banner, ROSMAP) (Fig. 7e-f and Supplementary Table 67). PWAS and TWAS identified 14 shared genes including *GSTT2B*, *SWAP70*, *HIBADH*, *ALDH2*, *NME7*, *HECTD4*, *LAMC1*, *ATG7*, *ICA1L*, *ACTR1B*, *SNX16*, *LYZ*, *FER* and *MIPEP*. Interestingly, *SWAP70* was a consistent signal across the TWAS, SMR, PWAS, and GWAS.

### Gene prioritization

We prioritized candidate genes for TIA by integrating GWAS, MR[MEGA, MTAG and downstream post[GWAS functional analyses. However, MR[MEGA and gene[based association analyses in stage 2 did not identified genome[wide significant signals, and thus were not contribute to the gene prioritization. The 17 lines of evidence actually used for gene prioritization included GWAS (stage 1, stage 2 and stage 3), MR-MEGA (stage 3), MTAG (stage 4), positional mapping (stage 1, stage 2 and stage 3), eQTL mapping (stage 1, stage 2 and stage 3), gene-based association analysis (stage 1 and stage 3), TWAS (stage 1), colocalization (stage 1), SMR (stage 1), and PWAS (stage 1). In total, 51 and 36 genes were supported by at least 4 and 5 lines of evidence (Fig. 4c, Supplementary Table 68). *SWAP70*, *KCNK3*, *SH2B3* and *CDKN2B* were robustly supported by at least 10 lines of evidence. Meanwhile, 3 genes (*CELSR2*, *NOS3*, *ALDH2*), 5 genes (*FOXF2*, *HDAC9*, *MMP3*, *ATXN2*, *HECTD4*), 5 genes (*TWIST1*, *MMP13*, *PSRC1*, *MAPKAPK5*, *NAA25*), and 6 genes (*PIK3CG*, *CDKN2B-AS1*, *COL4A1*, *MMP12*, *ACAD10*, *TMEM116*), were supported by 9, 8, 7 and 6 lines of evidence, respectively. Evidence showed that colocalization provided more conservative evidence than TWAS ^49^. In addition to the general gene prioritization by summarizing the different lines of evidence, we performed an additional gene prioritization focusing on genes with support from colocalization, PWAS, TWAS, and SMR. We highlighted *SWAP70* as a high-confidence gene, which showed consistent evidence across colocalization, PWAS, TWAS, and SMR (Supplementary Table 68).

### Drug-gene interaction analysis

To determine whether 51 TIA risk genes could serve as potential therapeutic targets, we utilized DGIdb 5.0 to explore the drug-gene interactions ^50^. In total, 24 of 51 TIA genes including *FGA*, *SH2B3*, *KCNK3*, *NOS3*, *MMP13*, *MMP12*, *TWIST1*, *CDKN2B*, *COL4A1*, *HDAC9*, and *ATXN2* showed evidence of interactions with 220 Food and Drug Administration (FDA)-approved drugs (Supplementary Table 69). *FGA* showed the strongest interactions with 9 FDA-approved drugs including Reteplase, Tirofiban, Eptifibatide, Plasminogen, Abciximab, Fibrinogen, Anistreplase, Plasmin, and Alteplase with interaction scores ranging from 0.48 to 4.77. The top 10 interactions included *FGA*-Reteplase (interaction score=4.77), *SH2B3*-Ruxolitinib (interaction score=4.63), *SLC4A1*-Metoprolol (interaction score=4.04), *PRKAR2B*-Theophylline (interaction score=3.89), *CUX2*-Rifampin (interaction score=3.75), *KCNK3*-Doxapram (interaction score=3.75), *SH2B3*-Candesartan (interaction score=3.50), *FGA*-Tirofiban (interaction score=1.90), *FGA*-Eptifibatide (interaction score=1.90), and *NOS3*-Arginine (interaction score=1.88).

### Differential gene expression analysis

We conducted a differential expression analysis of 51 TIA risk genes using gene expression data in brain and blood from 12 TIA mice at three time points 3h, 24h, and 72h after TIA (4 mice per mice), as well as 6 unhandled control mice (GEO accession: GSE32529) ^51,52^. All three post-TIA time points were compared with control mice for differential expression. In total, 46 of 51 TIA risk genes showed statistically significant differential expression including 11 in 3h brain, 18 in 3h blood, 28 in 24h brain, 28 in 24h blood, 8 in 72h brain, and 21 in 72h blood (|log_2_FC|≥0.2, FDR-adjusted *P*<0.05). Notably, 24h after TIA showed a relatively greater number of differentially expressed genes in both brain and blood. Specifically, 2 genes in brain (*Mmp12* and *Dcc*), and 10 genes in blood (*Swap70*, *Hdac9*, *Rasip1*, *Fes*, *Pmf1*, *Gosr2*, *Naa25*, *Ptpn11*, *Aldh2*, and *Lamc1*) were differentially expressed with |log_2_FC|≥1 and FDR-adjusted *P*<0.05. Interestingly, the top potential therapeutic targets *FGA* was upregulated in 72h blood (log_2_FC=0.44), *CUX2* was upregulated in 24h brain (log_2_FC=0.24) and 72h brain (log_2_FC=0.38), *KCNK3* was upregulated in 3h brain (log_2_FC=0.29), downregulated in 72h brain (log_2_FC=-0.29), and upregulated in 72h blood (log_2_FC=0.52), *NOS3* was upregulated in 24h brain (log_2_FC=0.59) and 24h blood (log_2_FC=0.39). Collectively, these findings demonstrate both shared and temporally dynamic transcriptional responses of TIA risk genes in the mouse model of TIA, with more significant expression changes (|log_2_FC|≥1) in blood than brain (Extended Fig. 3, Supplementary Tables 70-71).

### Tissue and cell analysis

To test potential enrichment of TIA heritability in specific human tissues or cells, we performed a linkage disequilibrium score regression for specific gene expression (LDSC-SEG) analysis ^53^ using stage 1 TIA GWAS meta-analysis summary data, GTEx v8 (49 tissues) and Franke laboratory multi-tissue expression data ^54,55^. However, no single tissue or cell passed the strict multiple-testing correction via the Bonferroni method (*P*<0.05/218). TIA heritability showed suggestive evidence of enrichment in 22 tissues, predominantly in brain regions including frontal lobe, hippocampus, entorhinal cortex, cerebral cortex, brain cortex, and arteries, and veins (*P*<0.05) (Supplementary Table 72).

We then focused on 51 TIA risk genes and evaluated their expression using GTEx (v10) gene expression data from 13 brain regions (amygdala, anterior cingulate cortex (BA24), caudate basal ganglia, cerebellar hemisphere, cerebellum, cortex, frontal cortex (BA9), hippocampus, hypothalamus, nucleus accumbens basal ganglia, putamen basal ganglia, spinal cord (cervical c-1) and substantia nigra), 3 blood vessles (aorta artery, coronary artery and tibial artery), 2 heart (heart atrial appendage and heart left ventricle), and whole blood. We revealed distinct tissue expression patterns among these genes. *RPL6*, *ERP29*, *PMF1*, *HIBAD*, *HPTPN11*, *TOM1L2*, *ALDH2* were widely expressed across almost all these tissues with relatively high transcript per million (TPM) values. *SLC4A1* was specifically expressed in blood. *COL4A1* and *SWAP70* displayed specifically high expression in blood vessels (aorta artery, coronary artery and tibial artery) (Extended Fig. 2d).

We further investigated the expression of 51 TIA risk genes using Human Brain Cell Atlas v1.0 single-nucleus RNA sequencing data from 606 high-quality samples covering 105 dissections across 10 brain regions obtained from four brains including three entire human brains from male donors and one motor cortex dissection from a female donor ^56^. We found widely expression of these risk genes in human brain nuclei, especially the top genes including *SWAP70* (central nervous system macrophage, endothelial cell, pericyte, vascular associated smooth muscle cell), *KCNK3* (pericyte, midbrain-derived inhibitory neuronal cell), *SH2B3* (central nervous system macrophage), *ALDH2* (astrocyte, bergmann glia, central nervous system macrophage, ependymal cell, endothelial cell, vascular associated smooth muscle cell), *CDKN2B* (choroid plexus epithelial cell, endothelial cell), *CELSR2* (oligodendrocyte), *HECTD4* (upper rhombic lip neuronal cells), *COL4A1* (vascular associated smooth muscle cell), *FOXF2* (pericyte, fibroblast, endothelial cell, vascular associated smooth muscle cell), and *HDAC9* (choroid plexus epithelial cell, midbrain-derived inhibitory neuronal cell, Extended Fig. 4).

## Discussion

Until now, most TIA genetic risk remains uncharacterized. Here, we performed three-stage single trait TIA GWAS and a cross-trait GWAS meta-analysis of TIA and stroke as well as its subtypes. Finally, we identified 44 independent loci including 25 known stroke loci and 19 TIA specific loci. Most loci were significantly associated with cerebrovascular diseases and risk factors, which provide critical insights into the genetic basis of TIA.

Among the known stroke loci, *CDKN2B-AS1* exhibited the most significant association with TIA in stage 1, stage 3 and stage 4, and was a known genome-wide significant locus for coronary artery disease ^57^, stroke, ischemic stroke, especially LAS ^13^. *CDKN2B-AS1* is a long non-coding RNA (lncRNA), and regulates the neighboring genes *CDKN2A* and *CDKN2B* that are cyclin-dependent kinase inhibitors ^58^. Loss of *CDKN2B* promotes atherosclerosis, and impairs TGF-β signaling and hypoxic neovessel maturation ^59,60^. The matrix metalloproteinase (MMP) family including *MMP3*, *MMP12* and *MMP13* were identified to be TIA loci. MMP are zinc-dependent enzymes crucial for extracellular matrix remodeling, inflammation, and tissue repair, with dual roles in maintaining homeostasis and mediating pathological injur<u>y</u> in neurovascular diseases ^61^. *MMP3* plays an important role in mediating cerebrovascular injury in hyperglycemic stroke ^62^. *MMP3* inhibition significantly reduced hemorrhagic transformation and improved functional outcomes in hyperglycemic stroke ^62^. Large-scale GWAS identified *MMP12* to be a genome-wide significant locus for stroke, ischemic stroke, especially LAS (*P*=4.39E-08) ^13^. *MMP13* is an important protease in infarct development and cortical remodeling during post-stroke neurorepair ^63^. Mendelian randomization showed that genetically predicted protein level of *MMP12* was associated with ischemic stroke ^64^. *PIK3CG* was associated with TIA in stage 1 (*P*=1.60E-09) and stage 3 (*P*=3.49E-11), and also was significantly associated with carotid atherosclerotic plaque (*P*=1.47E-10) ^65^, stroke, and ischemic stroke (*P*=4.94E-08) ^13^. *HDAC9* was associated with TIA in stage 1 (*P*=1.15E-10) and stage 3 (*P*=1.46E-13), and is a genome-wide significant locus for coronary artery disease ^57,66^, stroke, ischemic stroke, especially LAS (*P*=5.31E-13) ^13^. HDAC9-mediated activation of the NLRP3 inflammasome exacerbates chronic inflammation in mouse models of atherosclerosis ^67^. Meanwhile, large-scale GWAS have identified additional TIA loci as the genome-wide significant loci for stroke, ischemic stroke and coronary artery disease, including *KCNK3* (stroke, ischemic stroke) ^13^, *FGA* (stroke, ischemic stroke, and CES) ^13^, *FOXF2* (stroke, ischemic stroke) ^13^, *EVX1* (stroke)^13^, *NOS3* (stroke, ischemic stroke) ^13^, *SWAP70* (ischemic stroke, and coronary artery disease) ^13,57,66^, *SH2B3* (stroke, ischemic stroke) ^13^, *COL4A2* (stroke, ischemic stroke, SVS, and coronary artery disease) ^13,57,66^, *LDLR* (coronary artery disease, stroke and ischemic stroke) ^13,57,66^.

Most TIA specific loci are know genome-wide significant loci for blood pressures including *SLC4A7* (systolic blood pressure, diastolic blood pressure) ^68,69^, *CASC15* (systolic blood pressure, diastolic blood pressure, pulse pressure) ^69^, *GSE1* (systolic blood pressure) ^70^, *SLC4A1* (systolic blood pressure, pulse pressure) ^69^, *CELSR2* (diastolic blood pressure) ^71^. *CTAGE1*, *CELSR2* and *GOSR2* are genome-wide significant loci for coronary artery disease ^57,66,72,73^. *MRPS6* and *RBBP8* are genome-wide significant loci for burden of atherosclerosis ^72^, and intracranial aneurysm ^74^, respectively. *TUSC3* indicated the most significant DNA methylation in acute coronary syndrome ^75^.

Using gene set enrichment analysis, we highlighted statistically significant pathways, especially protein-lipid complexes, high-density lipoprotein particles, neurofibrillary tangles, and negative regulation of multicellular organismal processes. Lipids and lipoprotein particles crucially contribute to atherosclerosis, the underlying pathology of cardiovascular diseases ^76^. Observational literature search and Mendelian randomization analysis supported the involvement of negative regulation of multicellular organismal processes in peripheral artery disease ^77^. It is well known that neurofibrillary tangles, intracellular aggregates of hyperphosphorylated tau protein, are a fundamental neuropathological hallmark of Alzheimer’s disease ^78^. Meanwhile, the prevalence of pre-event dementia was 4.9% in patients with TIA, the incidence of post-event dementia at one year was 5.2% after TIA, and the risk of dementia at 1-5 years after TIA was 1.5 times higher ^79^.

Using a combination of 17 lines of evidence, we identified 51 genes supported by at least 4 lines of evidence (priority score≥4). Drug-gene interaction analysis further identified some of these genes as the potential therapeutic targets for TIA, especially the top interactions including *FGA*-Reteplase, *SH2B3*-Ruxolitinib, *PRKAR2B*-Theophylline, *CUX2*-Rifampin, *KCNK3*-Doxapram, and *SH2B3*-Candesartan. A randomized controlled trial showed that reteplase resulted in an excellent functional outcome than alteplase in ischemic stroke patients within 4.5 hours after symptom onset ^80^. Mendelian randomization demonstrated that genetically determined SH2B3 protein level was associated with coronary heart disease and ischemic stroke ^64^. Using middle cerebral artery occlusion mice, ruxolitinib treatment improved neurological scores, decreased the infarct size and ameliorated cerebral edema 3 days after stroke ^81^. Ruxolitinib ameliorated ischemic injury and neuroinflammation by reducing NLRP3 inflammasome activation via inhibiting JAK2/STAT3 pathway ^81^. Rifampin showed neuroprotective roles against global cerebral ischemia by activating Nrf2 pathway ^82^, and Alzheimer’s disease ^83^. Candesartan was developed to treat hypertension. In elderly patients with isolated systolic hypertension, candesartan treatment reduced 42% risk of stroke compared with other antihypertensive treatment ^84^. In Japanese hypertensive patients, especially in the patients with a past history of cardiovascular diseases, candesartan treatment was superior to other antihypertensive for reducing the risk of stroke and myocardial infarction ^85^. Until now, it remains unclear about the effects of these drugs on TIA patients, and further studies are warranted to determine whether these drugs are effective therapeutic agents for TIA.

Our current study may have several potential limitations. Firstly, the cross-ancestry GWAS meta-analysis was mainly based on European individuals from UKB, FinnGen, and MVP European. There were relatively small sample sizes in other non-European datasets including MVP AMR (2,792 cases and 55,391 controls) ^19^, MVP AFR (7,606 cases and 109,704 controls) ^19^, CKB (2,742 cases and 73,812 controls) ^26^, and GH (489 cases and 43,285 controls) ^28^. Secondly, only stage 1 TIA European-specific GWAS meta-analysis summary data was selected in multi[omic integrative analyses, such as TWAS and SMR, considering that most eQTL and pQTL datasets were from European individuals and the lack of mixed-ancestry LD scores. This strategy may contribute to reducing the bias from population stratification and reducing statistical power. Thirdly, the TIA GWAS summary statistics for X chromosome are publicly available in UKB and FinnGen but not in the other four cohorts including MVP, CKB, TWB and GH. On the one hand, the X chromosome is often excluded from GWAS due to its unique analytical challenges ^86^. On the other hand, the inclusion of the X chromosome, especially only in a subset of all cohorts, may not give sufficient statistical power, and the X chromosome data could not be directly compared with all autosomes from all cohorts. To maintain consistent and stable study cohorts, we excluded the X chromosome and focused on all autosomes from all cohorts. We acknowledge the important genetic contribution of the X chromosome, and plan to perform a TIA GWAS meta-analysis including the X chromosome when full X chromosome summary statistics are available from all included cohorts. Fourthly, genetic studies of TIA face fundamental challenges, particularly when relying on biobanks. However, the disease status collected in biobanks may not fully meet the strict diagnosis criteria, making it essential to replicate our findings using genetic data with phenotypes defined by diagnoses from neurological experts. Yet, until now, no publicly available TIA GWAS datasets use this case definition by neurologists. We will replicate our current findings when GWAS summary statistics from clinically diagnosed TIA cases become available. Fifthly, we confirmed the significant positive correlation between MVP and FinnGen TIA GWAS datasets, with *rg*=0.51 and *P=*9.99E-79. Compared with the UKB-FinnGen correlation (*r_g_*=0.92 and *P=*1.41E-02) and MVP-UKB correlation (*rg*=0.94 and *P=*8.20E-03), the MVP-FinnGen correlation was the most significant (*P=*9.99E-79) but not the strongest (*rg*=0.51). <u>This</u> <u>phenomenon may be</u> driven <u>by different</u> TIA <u>ICD-10 codes from different biobanks,</u> <u>including</u> ICD-10 code G45 in both UKB and FinnGen, and Phecode 433_31 in MVP. In fact, similar findings were also identified in recent GWAS meta-analyses. An anxiety GWAS meta-analysis identified significant positive correlations across different GWAS datasets ranging from 0.519 to 1 including the MVP-FinnGen <u>correlation</u> (*rg*=0.644 and *P=*1.56E-11), MVP-UKB <u>correlation</u> (*r_g_*=0.579 and *P=*1.67E-37), and UKB-FinnGen <u>correlation</u> (*rg*=0.866 and *P=*1.38E-15) ^87^. An epilepsy GWAS meta-analysis identified genetic correlations across datasets ranging from 0.31 to 0.74 ^88^. A thyroid GWAS meta-analysis highlighted genetic correlations across datasets ranging from 0.16 to 0.97 ^89^. Future studies are required to replicate our findings using the TIA GWAS datasets with the same <u>codes.</u> Taken together, our largest three-stage single trait TIA GWAS meta-analyses and cross-trait GWAS meta-analysis highlighted 44 genome-wide significant loci, 51 high-confidence genes, 13 statistically significant pathways, and 24 potential therapeutic targets for TIA. These genetic findings provide valuable insights into the underlying mechanisms of TIA, and broaden the potential therapeutic scope of the available drugs, which may contribute to future drug development targeting TIA.

## Methods

### Study participants

We selected 8 TIA GWAS datasets from UKB (European) ^17^, FinnGen (European) ^18^ (R12), MVP (European, AMR, and AFR) ^19^, CKB (East Asian) ^26^, TWB (East Asian) ^27^, and GH (South Asian) ^28^, with a total of 1,942,862 individuals including 82,533 TIA and 1,860,329 controls) (Supplementary Table 1). These prior studies received approval from the respective institutional review boards, and all enrolled participants gave written informed consent.

UKB is a population-based case-cohort prospective study that recruited over 500,000 participants aged 40-69 years between 2006 and 2010. UKB systematically integrates genome-wide genotyping data with longitudinal data from primary care, hospital admissions, cancer registries, death records, and participant self-reports, which allows researchers to cross-verify information, study the full spectrum of diseases, and build a robust, detailed health picture for over 500,000 participants, enabling powerful discoveries in medicine ^17^. UKB defined TIA as ’Transient cerebral ischaemic attacks’ using the International Classification of Diseases, Tenth Revision (ICD-10) code G45 including 3,150 cases and 417,381 controls of European ancestry.

FinnGen is a research project aimed at establishing a substantial Finnish cohort by integrating genetic information derived from biobanks with digital health record data obtained from health registries ^18^. FinnGen comes from the comprehensive national health registers covering the entire lifespan of the study subjects including hospital, diagnoses (ICD codes), special outpatient, primary care registers, drug prescription and causes of death, to build extensive genetic and clinical datasets for research ^18^. This data linkage is more completely than single data sources alone, provides unprecedented insight into conditions like TIA, and helps filter noise and find true cases for large-scale genetic studies. FinnGen defined TIA as ’Transient ischemic attack’ using ICD criteria (FinnGen code: I9, ICD-10 code: G45) including 24,948 cases and 453,276 controls of European ancestry in release 12 (Nov 4, 2024) ^18^.

MVP is an observational cohort study and mega-biobank in the Veterans Affairs health care system that collects data from participants via questionnaires, electronic health records, and blood samples, aiming to contribute to precision medicine ^19^. MVP uses sophisticated algorithms, often leveraging PheCodes (grouped ICD codes) that require specific patterns (multiple codes, specific dates) to boost accuracy, significantly raising the Positive Predictive Value for identifying conditions compared to just a single, standalone ICD code ^19^. MVP defined TIA as ’Transient cerebral ischemia (phecode 433_31)’ using an ICD code-based algorithm including 30,878 TIA and 402,820 controls of European ancestry, 2,792 cases and 55,391 controls of AMR ancestry, and 117,310 (7,606 TIA and 109,704 controls) of AFR.

CKB is a prospective cohort study of over 512,000 adults aged 30-79 years, recruited from 10 diverse regions across China between 2004 and 2008, focusing on exploring the lifestyle, environmental, and genetic determinants of common diseases ^26^. It gathered comprehensive baseline data and conducted additional measurements through resurveys involving approximately 5% of surviving participants. Disease follow-up is achieved through electronic linkage, utilizing unique national identity numbers to access death and disease registries, as well as the Chinese national health insurance system ^26^. CKB defined TIA using ICD-10 code G45 including 2,742 cases and 73,812 controls ^26^.

TWB is a prospective study of over 150,000 Taiwanese aged 20-70, collecting comprehensive phenotypic and genetic data, emphasizing genetic quality, population structure (Han Chinese), and findings, with ongoing linkage to health databases for deeper research ^27^. TWB defined TIA as ’Transient cerebral ischemia (433.31)’ using ICD-10 code G45 including 9,928 cases and 304,660 controls ^27^.

GH is a community-based population genomics and health initiative that focuses on approximately 50,000 British individuals of South Asian ancestry (Bangladeshi and British Pakistani) recruited in the United Kingdom, with contributions from regions including East London (East London Genes & Health program) and Bradford (Bradford Genes & Health initiative) ^28^. GH diagnosed TIA using ICD-10 including 489 cases and 43,285 controls ^28^.

### Single GWAS level quality control

Before meta-analysis, rigorous quality control was applied to each GWAS dataset. All GWAS datasets were processed to obtain a harmonized format. We converted the build 38-imputed genotype genome coordinates from FinnGen and MVP to their corresponding build 37 (hg19) coordinates using CrossMap ^90^ and Python 3.6. Using the 1000 Genomes Phase 3 dataset as a reference ^91^, we performed a genetic variant quality control to exclud variants: (1) non-standard alleles (other than A, T, C, G); (2) monomorphic SNPs; (3) duplicated or multiallelic SNPs; (4) SNPs absent from the reference and SNPs located on the X or Y chromosome ^92,93^; (5) SNPs with a MAF<1%. Consequently, only biallelic SNPs with ≥1% were included in meta-analysis. The total number of the excluded SNPs is provided in Supplementary Table 72.

### GWAS meta-analysis and quality control

After the quality control, TIA GWAS meta-analysis was performed using a fixed-effects inverse variance weighted method implemented by METAL (v 2011-03-25), weighted by the effect size and stander error from each genetic variant, with genomic control correction ^21^. Here, we performed three-stage TIA GWAS meta-analyses including stage 1 European-specific meta-analysis (UKB, FinnGen, and MVP European, 1,332,453 individuals including 58,976 cases and 1,273,477 controls), stage 2 non-European meta-analysis (MVP AMR, MVP AFR, CKB, TWB, GH, 610,409 individuals including 23,557 cases and 586,852 controls, and stage 3 cross-ancestry meta-analysis (UKB, FinnGen, MVP European, MVP AMR, MVP AFR, CKB, TWB and GH, 1,942,862 individuals including 82,533 cases and 1,860,329 controls).

In each stage, we filtered the GWAS meta-analysis results to exclude genetic variants present in <50% of the included GWAS datasets (e.g., fewer than 2 of 3 datasets in stage 1, fewer than 3 of 5 datasets in stage 2 and fewer than 4 of 8 datasets in stage 3). In stage 3 cross-ancestry meta-analysis, we only selected genetic variants present in at least one European and at least one non-European population cohort to ensure the validity of the cross-ancestry analysis. Genetic variants were considered significant using the genome-wide significance level (*P*<5.00E-08).

We further calculate genomic inflation factor (λ_GC_) and LDSC intercept using LDSC (v1.0.1) and LD scores from the 1000 Genomes Project phase 3 European reference panel ^20^. LDSC estimates the liability-scaled heritability for TIA using the observed-scale heritability from GWAS summary statistics and adjusting it using the population prevalence and sample prevalence, providing a standardized measure of genetic contribution to disease risk ^20^. If the population prevalence is not available, a lifetime risk estimate is usually used. It was estimated that the population prevalence of TIA was 2.3% in the United States ^22^. Here, we calculated the genome-wide SNP-based heritability on the liability scale using LDSC (v1.0.1) assuming the TIA population prevalence 2.3% ^22^.

In stage 2 and stage 3, we conducted a sensitivity analysis using a multi-ancestry meta-analysis implemented in MR-MEGA (v0.2) to account for potential heterogeneity ^29^. MR-MEGA detects complex trait association signals via multi-ancestry meta-regression ^29^. MR-MEGA uses genome-wide allele frequency differences to create Principal Components (PCs) (axes of genetic variation) via Multi-Dimensional Scaling, then includes these PCs in a linear regression model to meta-analyze GWAS data, effectively accounting for population structure and improving detection of variants with ancestry-correlated effects, boosting power and fine-mapping resolution in diverse populations ^29^. MR-MEGA enables partitioning of the heterogeneity into components due to ancestry and residual variation, quantified by *P*_value_ancestry_het_ and *P*_value_residual_het_, respectively ^29^.

### Identification of genetic risk loci

TIA risk loci were identified using FUMA (v.1.5.2) by default parameters ^23^. Initially, FUMA identified SNPs with the genome-wide significance of *P<*5.00E-08 and LD threshold *r*^2^<0.6 as the independent significant SNPs using LD-based clumping ^23^. The independent genetic risk loci were characterized by considering all SNPs in LD (*r*^2^≥0.6) with one of the independent significant SNPs within a region of 250 kilobases (kb) ^23^. Within each genetic risk locus, FUMA further distinguished the lead SNPs, which are a subset of the independent significant SNPs in LD with each other (*r*^2^<0.1) ^23^. Each locus was represented by the lead SNP with the most significant *P* value. LD information is from the 1000 Genomes Project phase 3 European panel (stage 1) and the all-ancestry panel (stage 2, stage 3 and stage 4) ^23^.

### Genetic association analysis

Firstly, before stage 1 GWAS meta-analysis, we evaluated the pairwise genetic correlation across the three TIA GWAS datasets from UKB, FinnGen, and MVP European using LDSC (v1.0.1) with the default settings and LD information from the 1000 Genomes Project phase 3 European reference panel ^23^. Here, we did not conduct a cross-ancestry genetic correlation analysis of TIA GWAS datasets as the existing GWAS summary statistic methods remain unreliable for admixed populations ^94,95^. Secondly, after stage 1 GWAS meta-analysis, we estimated the pairwise genetic correlation (*rg*) of TIA (stage 1 European) between stroke, ischemic stroke and its subtypes including LAS, CES and SVS. Analyses were performed in LDSC (v1.0.1) with default settings, using large-scale GWAS datasets from the MEGASTROKE consortium (European ancestry) ^30^, to evaluate the potential of combining TIA and stroke in cross-trait GWAS meta-analysis. Here, we selected the LD information from the 1000 Genomes Project Phase 3 European reference panel, as the TIA and stroke GWAS datasets were from individuals of European ancestry ^39^. Thirdly, we evaluated the genetic association of TIA loci from four stages with stroke and its subtypes using large-scale GWAS datasets from the MEGASTROKE and GIGASTROKE consortia ^13^.

### Cross-trait GWAS meta-analysis of TIA and stroke

We conducted a cross-trait GWAS meta-analysis of TIA with stroke and its subtypes including ischemic stroke, LAS, CES and SVS using MTAG (v1.0.8) with default settings ^31^. MTAG jointly analyzes GWAS summary statistics from multiple genetically correlated traits, improves the power for discovery of genetic loci for each trait and explores the shared genetic etiology by boosting the effective sample size ^31^. It is noted that MTAG was developed to perform a meta-analysis in individuals of the same ancestry, as cross-ancestry MTAG violates the assumptions of the model ^31^. Here, MTAG was performed in individuals of European ancestry using stage 1 TIA GWAS meta-analysis and GWAS datasets for stroke and its subtypes. To reduce bias caused by sample overlap, we selected GWAS datasets for stroke and its subtypes from the MEGASTROKE consortium (excluding UKB and FinnGen) ^30^ rather than from the GIGASTROKE consortium (including UKB and FinnGen) ^13^.

### Phenome-wide association study

We performed a PheWAS to improve the understanding of the clinical phenotypes associated with the GWAS significant TIA genetic variants. In breif, we evaluated the genetic association of TIA genetic variants with a broad range of phenotypes using three online PheWAS resources including UKB PheWeb (1,400 traits, 57 million TOPMed-imputed variants in 400,000 British white individuals) ^32^, MVP (2,068 traits, 449,042 Europeans) ^19^ and FinnGen R12 (2,469 traits, 500,348 individuals) ^33^. To ensure the statistical power, phecodes with fewer than 200 cases were excluded. Bonferroni correction was applied to account for multiple testing across 5,937 traits with *P*<0.05/5937*=*8.42E-06.

### Gene mapping

We assigned independent significant SNPs and their proxy SNPs (*r*^2^≥0.6) to specific genes using positional mapping and eQTL mapping. Positional mapping is performed using ANNOVAR implemented in FUMA with default settings, by mapping independent significant SNPs and their proxy SNPs (*r*^2^≥0.6) to protein-coding genes within a 10 kb window ^23^. eQTL mapping aims to identify genes whose expression levels in specific tissues are regulated by the independent significant SNPs and their proxy SNPs (*r*^2^≥0.6) within a range of 1 megabase (Mb) (*cis*-eQTLs) using well established eQTL datasets. Here, we selected eQTL data from GTEx v10, which primarily consists of individuals of European ancestry, including 13 brain tissues (amygdala, anterior cingulate cortex (BA24), caudate basal ganglia, cerebellar hemisphere, cerebellum, cortex, frontal cortex (BA9), hippocampus, hypothalamus, nucleus accumbens basal ganglia, putamen basal ganglia, spinal cord (cervical c-1) and substantia nigra), 3 blood vessels (aorta artery, coronary artery and tibial artery), 2 heart tissues (heart atrial appendage and heart left ventricle), and whole blood from 838 postmortem donors ^24^. We also included brain eQTL data from MetaBrain, a large-scale meta-analysis comprising of 2,683 individuals of European ancestry ^25^. An FDR-corrected *P*<0.05 was considered statistically significant for eQTL mapping.

### Gene-based association study and gene set enrichment analysis

We conducted a gene-based association study and gene set enrichment analysis of TIA GWAS meta-analysis data using MAGMA (v1.10). MAGMA used a multiple regression model for gene analysis and a competitive approach for gene-set analysis, respectively ^34^. MAGMA maps all SNPs from TIA GWAS meta-analysis to their corresponding genes through human reference genome build 37 genomic coordinates and LD information from the 1000 Genomes Project phase 3 European reference panel, and provides a gene-based association score by the aggregate of all SNPs inside each gene by accounting for gene size, variant density and LD structure ^34^. Here, we selected the European reference panel, as the majority of GWAS cohorts in our study were of European ancestry, making it the most statistically appropriate and powerful reference ^96^. We focused on gene sets from GO terms including biological processes, cellular components and molecular functions from the Molecular Signatures Database (MSigDB) (version 2024.1.Hs). An FDR-corrected *P*<0.05 was considered statistically significant for both gene-based association test and gene set enrichment analysis.

### Transcriptome-wide association study

TWAS is an effective approach to identify novel susceptibility genes by systematically investigating the association of genetically predicted gene expression with disease risk. In brief, TWAS integrates GWAS with eQTL data to find genes whose expression levels, driven by genetic variants, are link to complex traits or diseases, offering deeper biological insights than GWAS alone. Here we conducted a TWAS implemented by FUSION with default settings ^38^, by integrating stage 1 TIA GWAS meta-analysis data with eQTL data from CMC brain (537 individuals in dorsolateral prefrontal cortex) ^35^ and GTEx v8 ^24^, including 13 brain tissues (amygdala, anterior cingulate cortex (BA24), caudate basal ganglia, cerebellar hemisphere, cerebellum, cortex, frontal cortex (BA9), hippocampus, hypothalamus, nucleus accumbens basal ganglia, putamen basal ganglia, spinal cord (cervical c-1) and substantia nigra), 3 blood vessels (aorta artery, coronary artery and tibial artery), 2 heart tissues (heart atrial appendage and heart left ventricle), and whole blood. An FDR-corrected *P*<0.05 was considered statistically significant.

### Colocalization analysis

We conducted a genetic colocalization analysis of TWAS significant signals (FDR<0.05) using COLOC implemented in FUSION ^38^ to investigate whether TIA GWAS and eQTL data from CMC ^35^ and GTEx v8 ^24^ share the same single causal genetic variant in a genomic region. COLOC tests hierarchical hypotheses to find the posterior probability (PP) of colocalized association: no association with either trait (PPH0), association with only one trait (PPH1, PPH2), association with both traits but different causal variants (PPH3), association with both traits sharing the same single causal variant (PPH4) ^38^. A PPH4 ≥0.8 indicated the strong evidence for colocalization ^38^.

### Summary-data-based mendelian randomization

Both TWAS and SMR aim to identify novel disease risk genes by assessing the relationship between disease and the expression of specific genes ^43^. TWAS could not determine whether genes have the causal effects on disease risk ^42^. SMR utilizes a Mendelian randomization method to determine the causal effects of gene expression on disease risk ^43^. Here, we conducted SMR using SMR (v1.3.1) with default settings, integrating the stage 1 TIA GWAS meta-analysis summary data with eQTL data from GTEx v8 ^24^ (13 brain tissues including amygdala, anterior cingulate cortex BA24, caudate basal ganglia, cerebellar hemisphere, cerebellum, cortex, frontal cortex BA9, hippocampus, hypothalamus, nucleus accumbens basal ganglia, putamen basal ganglia, spinal cord cervical c-1 and substantia nigra), large blood eQTL dataset from eQTLGen (31,684 individuals) ^44^, and brain eQTL dataset from BrainMeta v1 (a maximum of 1,194 samples) ^45^. An FDR-corrected *P*<0.05 and a heterogeneity *P*>0.01 from the HEIDI test were considered statistically significant.

### Proteome-wide association study

PWAS is a natural extension of TWAS approach by integrating pQTL data with GWAS data to evaluate the associations of genetically predicted protein levels and disease ^38^. The main difference between TWAS and PWAS is the molecular layer: TWAS links genetics to gene expression levels (mRNA) in specific tissues, while PWAS links genetics to protein abundance (like in blood and brain), with both aiming to identify genes underlying complex traits using genetic data (GWAS) ^38^. To discover genes whose protein levels are linked to TIA, we conducted a PWAS implemented in FUSION ^38^ by combining stage 1 TIA GWAS meta-analysis summary data with two brain pQTL datasets from Banner (dorsolateral prefrontal cortex, 152 individuals of European ancestry across 8,168 proteins) and ROS/MAP (dorsolateral prefrontal cortex, 376 participants of European ancestry across 8,356 proteins) ^46,47^, and a blood pQTL dataset from Atherosclerosis Risk in Communities (ARIC) study (plasma, 7,213 individuals of European ancestry) ^48^. It is noted that both the stage 1 TIA GWAS and pQTLs were of European ancestry. To be consistent with TWAS, we selected the LD information from the 1000 Genomes Project Phase 3 European reference panel in PWAS, given that all pQTL datasets were of European ancestry. The proteins with FDR<0.05 were considered statistically significant. Accordingly, the genes corresponding to these proteins were defined as the TIA risk genes.

### Gene prioritization

To identify the most probable causal genes for TIA, we integrated multiple lines of evidence from different analysis methods including GWAS meta-analysis using METAL (stage 1, stage 2 and stage 3), cross-ancestry meta-analysis using MR-MEGA (*P*<5.00E-08, stage 2 and stage 3), cross-trait meta-analysis using MTAG (*P*<5.00E-08, stage 4), positional mapping (FUMA, stage 1, stage 2 and stage 3), eQTL mapping (FDR<0.05, stage 1, stage 2 and stage 3), gene-based association analysis using MAGMA (FDR<0.05, stage 1, stage 2 and stage 3), TWAS (FDR<0.05, stage 1), colocalization (FDR<0.05, stage 1), SMR (FDR<0.05, stage 1), and PWAS (FDR<0.05, stage 1). For each gene, the prioritization score is the sum of multiple multiple lines of evidence by counting “0” if the gene was ‘not prioritized’ and as “1” if the gene was ‘prioritized’. This approach ensured that genes with stronger cumulative evidence were more likely to be causally associated with TIA, and reflected a higher degree of confidence in the TIA etiology.

### Drug-gene interaction analysis

To determine whether the TIA risk genes could serve as potential therapeutic targets, we utilized DGIdb 5.0 to explore the drug-gene interactions by searching the candidate genes against the known and potentially druggable genome ^50^. A drug-gene interaction represents a documented relationship between a gene (or its protein product) and a pharmaceutical compound, curated from multiple sources including scientific literature, databases, and clinical annotations ^50^. These relationships encompass various biological mechanisms, including direct physical binding (e.g., drug-target interactions); pharmacogenomic associations (e.g., genetic variants affecting drug response); and functional modulation (e.g., activator, inhibitor, or modulator relationships) ^50^. DGIdb calculated an interaction score to rank the interaction results, where higher scores indicate stronger evidence supporting drug-gene interactions ^50^.

### Differential gene expression analysis

We evaluated the differential expression of TIA risk genes using gene expression dataset from the mouse model of TIA (GEO accession: GSE32529) ^51,52^. Brain ipsilateral cortex tissue and blood were collected and processed from 6 unhandled control mice, and 12 TIA mice at three time points including 3h, 24h, and 72h after TIA (4 mice per group) ^51,52^. Differential expression analysis between each TIA group and control group was performed using GEO2R tool, and genes with |log_2_FC|≥0.2 and FDR-adjusted *P*<0.05 were considered statistically significant.

### Tissue and cell enrichment analysis

In order to investigate whether TIA heritability is enriched in specific tissues or cells, we performed a tissue and cell enrichment analysis using LDSC-SEG ^53^ to integrate the stage 1 TIA GWAS meta-analysis summary statistics with eQTL datasets from GTEx v8 ^24^ (49 tissues) and a multi-tissue gene expression dataset from the Franke laboratory (37,427 human samples from DEPICT website) ^54,55^. The original *P* values were adjusted using the Bonferroni method to account for multiple testing (*P*[<[0.05/218). Tissue-specific expression of TIA risk genes were visualized using the GTEx Portal (v10) multi-gene query tool. Median transcripts per million (TPM) values were extracted for each gene across brain, blood, vasculature, and heart. We further examined cell-specific expression of TIA risk genes using Human Brain Cell Atlas v1.0 implemented by Human Protein Atlas (HPA), which included single-nucleus RNA sequencing data from 606 high-quality samples, 105 dissections, and 10 brain regions obtained from four brains including three entire human brains from male donors and one motor cortex dissection from a female donor ^56^. A graph-based clustering hierarchically divided 3,369,219 cells into 31 superclusters, 461 clusters, and 3313 subclusters ^56^. A normalized single nuclei RNA nTPM value was used to evaluate the expression from each gene.

## Data availability

TIA GWAS summary statistics is available at figshare, https://figshare.com/s/394de134305092d00431. The GWAS summary statistics generated in this study were obtained from the following sources: UKB, https://pan.ukbb.broadinstitute.org/; FinnGen, https://r12.finngen.fi/; MVP, https://www.ncbi.nlm.nih.gov/projects/gap/; CKB, https://pheweb.ckbiobank.org/phenotypes; British South Asian (Genes & Health GWAS), https://www.genesandhealth.org/research/gwas-data-downloads; TWB, https://pheweb.ibms.sinica.edu.tw/. The LD scores are from the 1000 genomes phase 3 European reference panel are available at https://data.broadinstitute.org/alkesgroup/. The phenotype information were obtained from the following sources: UKB, https://pheweb.org/UKB-TOPMed/; MVP, https://phenomics.va.ornl.gov/web/cipher/pheweb; FinnGen, https://r12.finngen.fi/coding; CKB, https://pheweb.ckbiobank.org/; TWB, https://pheweb.ibms.sinica.edu.tw/. The SNP-expression weight files are available at http://gusevlab.org/projects/fusion/. The SNP-protein weight files are available at https://www.synapse.org/ (Synapse ID: syn23191787) and http://nilanjanchatterjeelab.org/pwas/. The eQTL data for SMR are available at https://yanglab.westlake.edu.cn/software/smr/#DataResource. Franke lab’s multi-tissue expression data are available at https://data.broadinstitute.org/mpg/depict/depict_download/tissue_expression. DGIdb is available at https://dgidb.org.

## Code availability

No custom code was used in this study. As detailed in the Methods section, we used previously released software to generate the analysis and cite them throughout the manuscript references. FUMA (v.1.5.2) is available at https://fuma.ctglab.nl/. METAL (v 2011-03-25) is available at https://csg.sph.umich.edu/abecasis/Metal/. MR-MEGA (v0.2) is available at https://genomics.ut.ee/en/tools. MTAG (v1.0.8) is available at https://github.com/JonJala/mtag. LocusZoom is available at http://locuszoom.sph.umich.edu/. LDSC (v1.0.1) is available at https://github.com/bulik/ldsc. MAGMA (v1.10) is available at https://cncr.nl/research/magma/. FUSION http://gusevlab.org/projects/fusion/. SMR is is available available at at https://yanglab.westlake.edu.cn/software/smr/#SMR. LDSC-SEG is available at https://github.com/bulik/ldsc/wiki/Cell-type-specific-analyses.

## Acknowledgments

This work was supported by funding from the National Key Research and Development Program of China (Grant No. 2023YFC3605200), National Natural Science Foundation of China (Grant No. 82471449, 82071212, and 81901181), Beijing Natural Science Foundation (Grant No. JQ21022). We acknowledge the contribution of participants and investigators of the UK Biobank, FinnGen, Million Veteran Program, China Kadoorie Biobank, Genes and Health study and Taiwan Biobank, contributing to the GWAS summary statistics in our analysis.

## Author contributions

Guiyou Liu and Xunming Ji conceived the study. Guiyou Liu and Shuyuan Hu designed and supervised the analyses. Shuyuan Hu carried out the analyses with support from Ping Zhu, Shiyang Wu and Shan Gao. Shan Gao, Fengzhen Liu and Yijie He assisted with data visualization. Zhifa Han, Tao Wang, Mingxin Wang, Changhong Ren, Wenbo Zhao and Sijie Li advised on the study. Shuyuan Hu wrote the manuscript with input and support from all authors. All authors contributed to the interpretation of the results and critical revision of the manuscript and approved the final version of the manuscript.

## Ethics approval and consent to participate

This article contains human participants collected by several studies performed by previous studies. All participants gave informed consent in all the corresponding original studies. Here, our study is based on the publicly available, large-scale datasets, and not the individual-level data. Hence, ethical approval was not sought.

## Consent for publication

All authors give consent for publication.

**Extended Fig. 1 Locuszoom plots of 8 genome-wide significant TIA loci from Non-European-specific meta-analysis (stage 2).**

LocusZoom plots of rs9856538 (*SLC4A7*), rs201920009 (*CASC15*), rs6970288 (*SRRM3*), rs73159770 (*NOS3*), rs10991625 (*SLC44A1*), rs76670928 (*LOC107984361*), rs12923257 (*GSE1*), rs17225358 (*LOC105372530*) The *x* axis depicts the chromosome and base pair positions of genetic variants. The *y* axis shows the -log_10_(*P* values). *P* values are from the fixed-effects inverse variance weighted method implemented by METAL. All statistical tests were two-sided. The grey dash line shows the genome-wide significance threshold (*P*<5.00E-08).

**Extended Fig. 2 Pathway enrichment analysis across three stages and GTEx tissue-specific expression of TIA prioritized risk genes.**

Gene set enrichment analysis of TIA GWAS meta-analysis data using MAGMA in (a) Stage 1, (b) Stage 2 and (c) Stage 3. The significance level accounts for multiple testing of gene sets, utilizing a false discovery rate (FDR)-corrected threshold of *P*<0.05. The Figure shows top 10 pathways with -log_10_(FDR) and the significant pathways are marked in red. BP, biological process; CC, cellular component; MF, molecular function. (d) Heatmap showing transcript per million (TPM) expression levels of the associated genes across brain, heart, blood vessels and whole blood tissues from GTEx. Colour scale ranges from white (TPM=0) to dark blue (TPM≥15), with tissues labelled along the bottom x-axis and genes along the right y-axis.

**Extended Fig. 3 Differential expression analysis of TIA prioritized risk genes in mouse brain and blood.**

Volcano plots showing differential expression of the 51 TIA risk genes in mouse brain at (a) 3h, (b) 24h, (c)72h and in blood at (d) 3h, (e) 24h, (f) 72h after TIA, compared with unhandled controls (n=4 TIA mice per time point; n=6 controls). The *x* axis represents logL(fold change) and the y-axis represents -log_10_(*P*-value). Blue dots indicate significantly downregulated genes (TIA low), red dots indicate significantly upregulated genes (TIA high), and gray dots represent non-significant genes. (g) Summary of significantly differentially expressed genes across all conditions.

**Extended Fig. 4 Cell type-specific expression of TIA prioritized risk genes in Human Brain Cell Atlas v1.0 from Human Protein Atlas (HPA).**

Bar plots illustrating the cell-type expression of 51 TIA risk genes using Human Brain Cell Atlas v1.0 single-nucleus RNA sequencing data from 606 high-quality samples covering 105 dissections across 10 brain regions obtained from four brains including three entire human brains from male donors and one motor cortex dissection from a female donor, as implemented by Human Protein Atlas (HPA). Panels (a-j) correspond to the top 10 genes including *SWAP70*, *KCNK3*, *SH2B3*, *ALDH2*, *CDKN2B*, *CELSR2*, *HECTD4*, *MMP13*, *COL4A1*, and *FOXF2*. A normalized single nuclei RNA nTPM value was used to evaluate the expression from each gene. Color-coding is based on cell type groups (as defined in the single cell type data representing the whole body), in this case separated into the main cell categories of neuron, glial cells, endothelial cells, fibroblasts and muscle cell (vascular smooth muscle cells and pericytes).

