## Supplementary Figures for "Genome-wide association study highlights 44 loci for transient ischemic attack and shared genetic architecture with stroke"

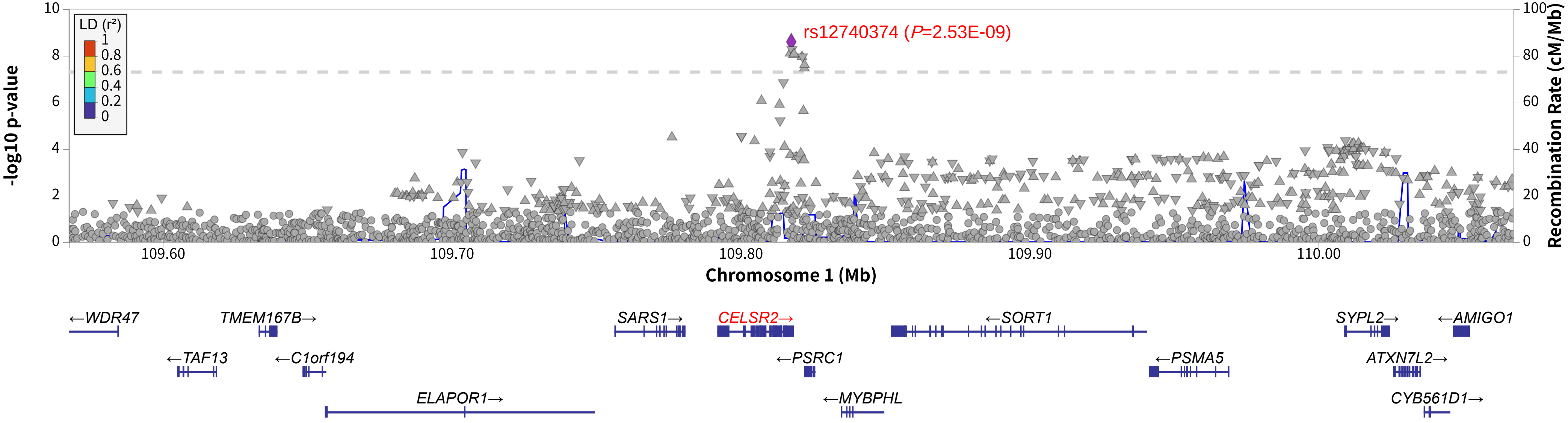


**Supplementary Fig. 1 Locuszoom plots for rs12740374 of cross-ancestry meta-analysis (stage 3, METAL) genome-wide significant association.** rs12740374is located downstream of *CELSR2*. The *P*-values are from fixed-effects model GWAS meta-analyses, and all association analyses were two-sided.

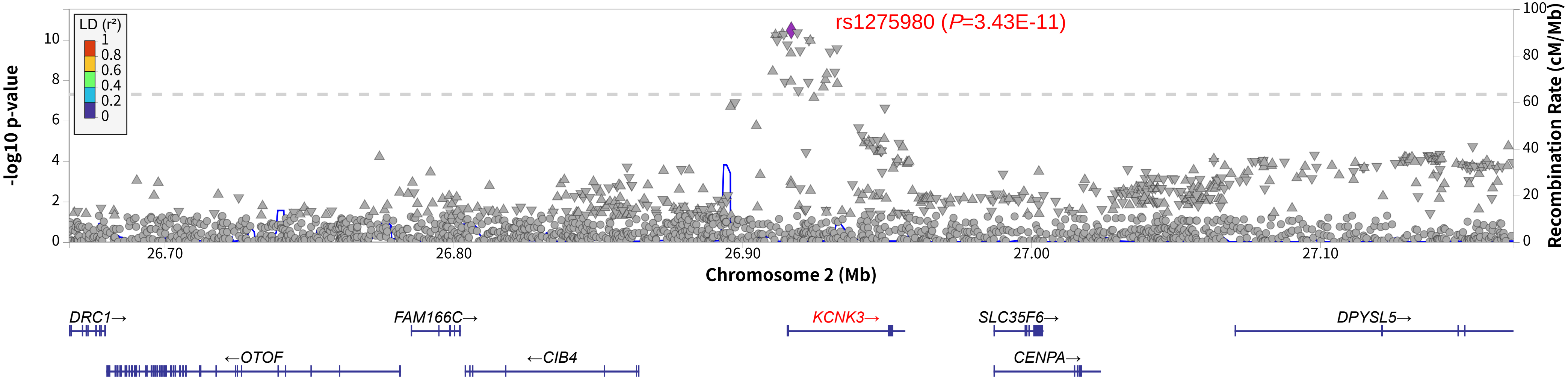


**Supplementary Fig. 2 Locuszoom plots for rs1275980 of cross-ancestry meta-analysis (stage 3, METAL) genome-wide significant association.** rs1275980is located on *KCNK3.* The *P*-values are from fixed-effects model GWAS meta-analyses, and all association analyses were two-sided.

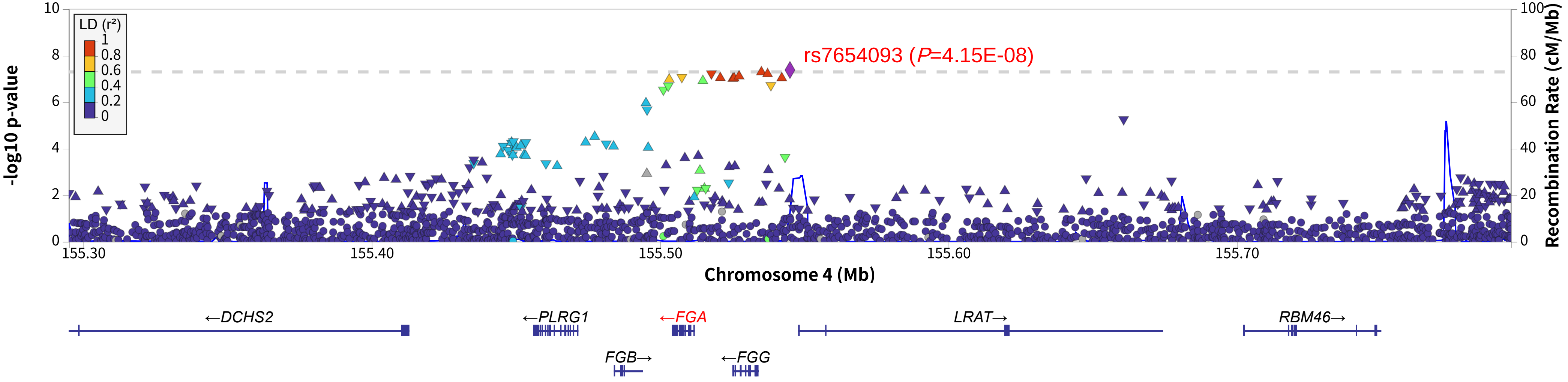


**Supplementary Fig. 3 Locuszoom plots for rs7654093 of cross-ancestry meta-analysis (stage 3, METAL) genome-wide significant association.** rs7654093is located upstream of *FGA.* The *P*-values are from fixed-effects model GWAS meta-analyses, and all association analyses were two-sided.

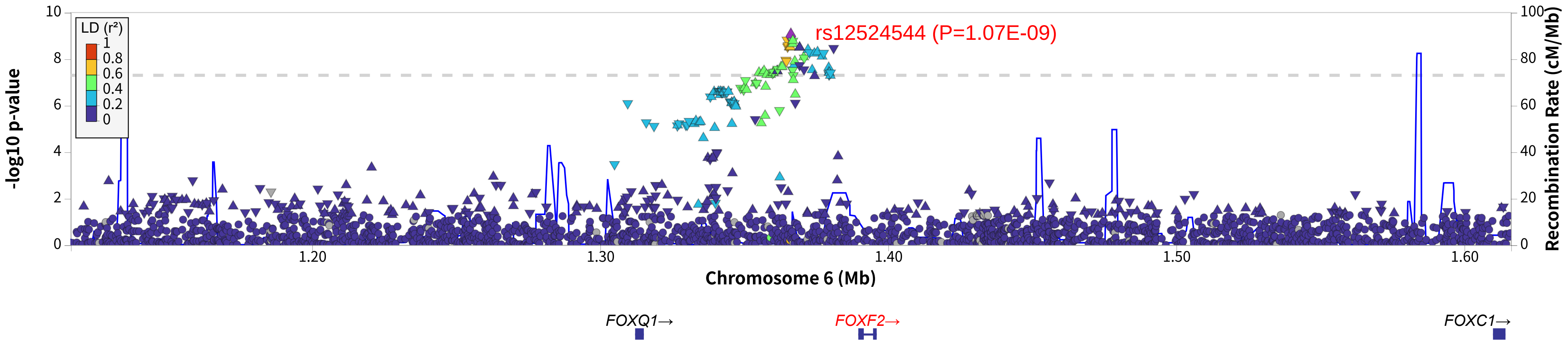


**Supplementary Fig. 4 Locuszoom plots for rs12524544 of cross-ancestry meta-analysis (stage 3, METAL) genome-wide significant association.** rs12524544 is located upstream of *FOXF2*. The *P*-values are from fixed-effects model GWAS meta-analyses, and all association analyses were two-sided.

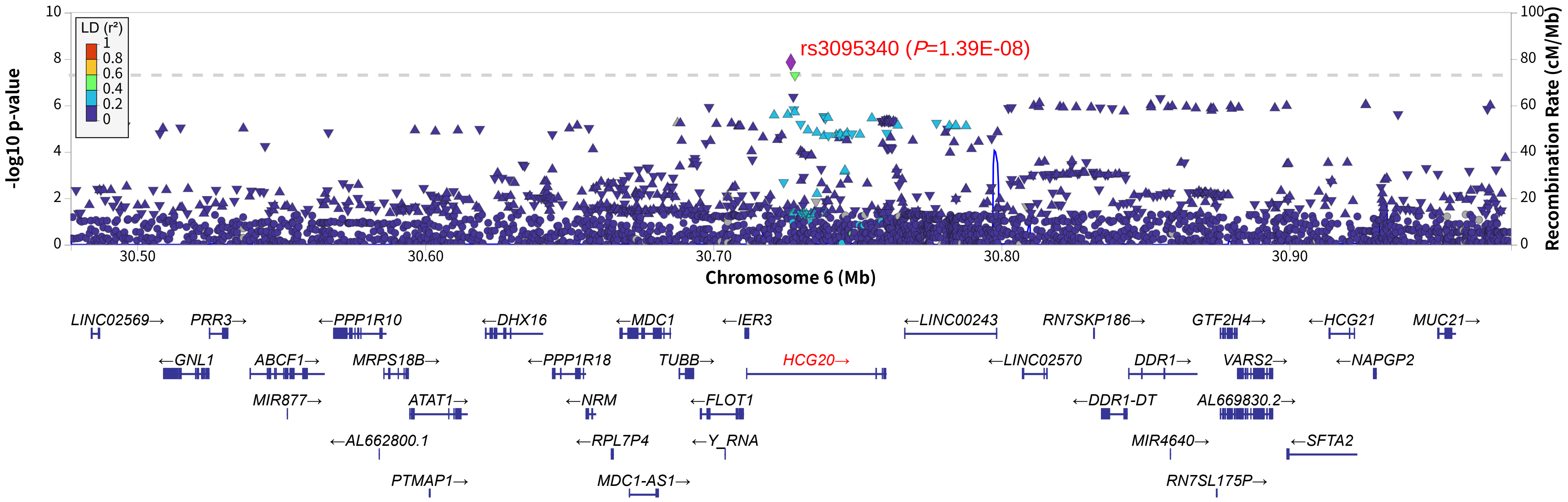


**Supplementary Fig. 5 Locuszoom plots for rs3095340 of cross-ancestry meta-analysis (stage 3, METAL) genome-wide significant association.** rs3095340 is located on *HCG20.* The *P*-values are from fixed-effects model GWAS meta-analyses, and all association analyses were two-sided.

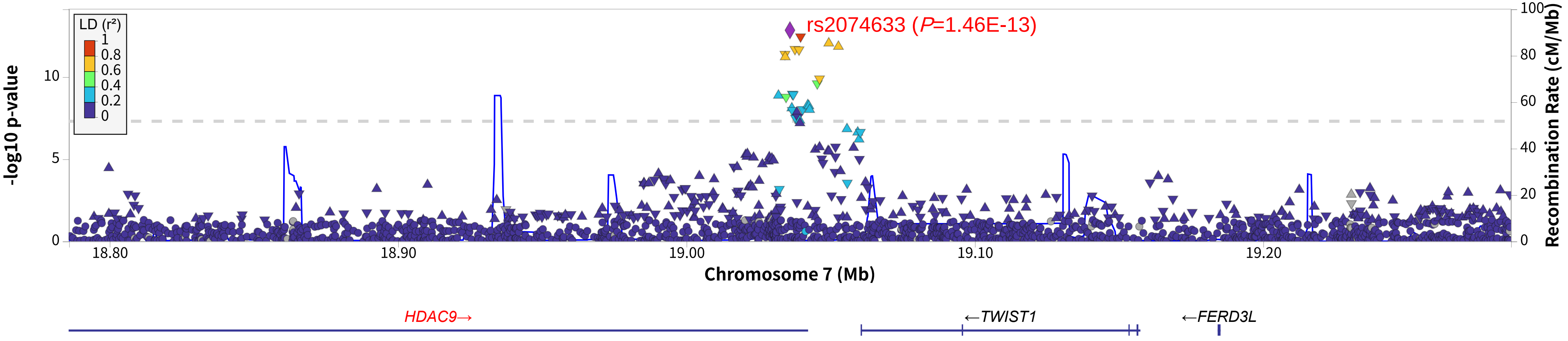


**Supplementary Fig. 6 Locuszoom plots for rs2074633 of cross-ancestry meta-analysis (stage 3, METAL) genome-wide significant association.** rs2074633 is located on *HDAC9.* The *P*-values are from fixed-effects model GWAS meta-analyses, and all association analyses were two-sided.

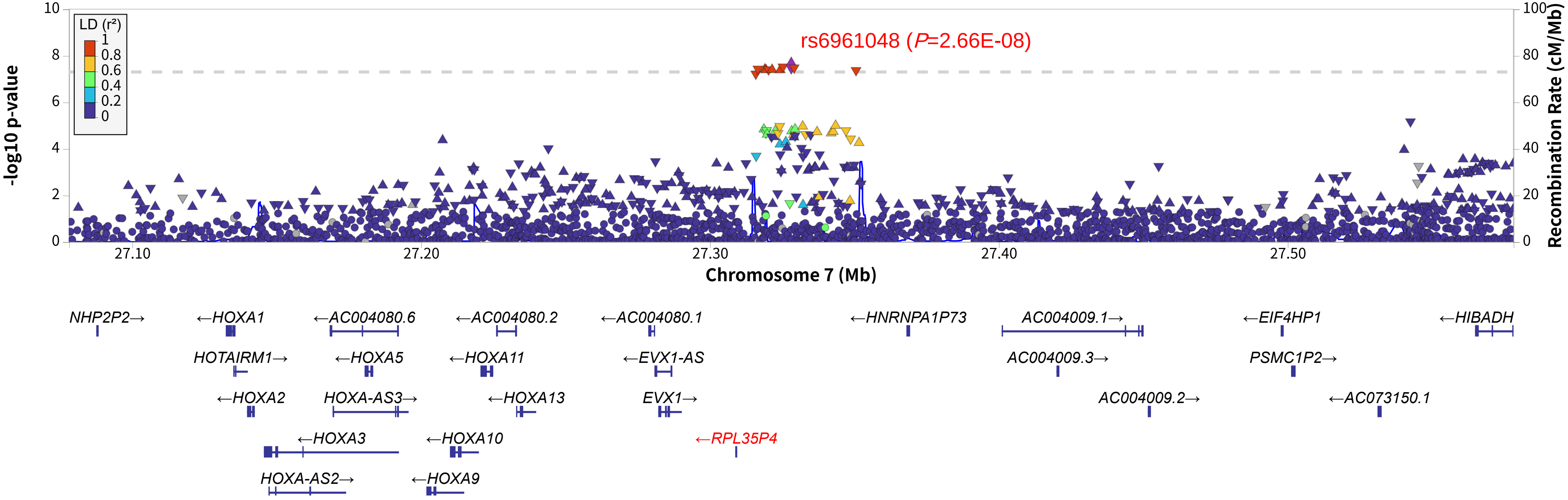


**Supplementary Fig. 7 Locuszoom plots for rs6961048 of cross-ancestry meta-analysis (stage 3, METAL) genome-wide significant association.** rs6961048is located upstream of *RPL35P4*.The *P*-values are from fixed-effects model GWAS meta-analyses,and all association analyses were two-sided.

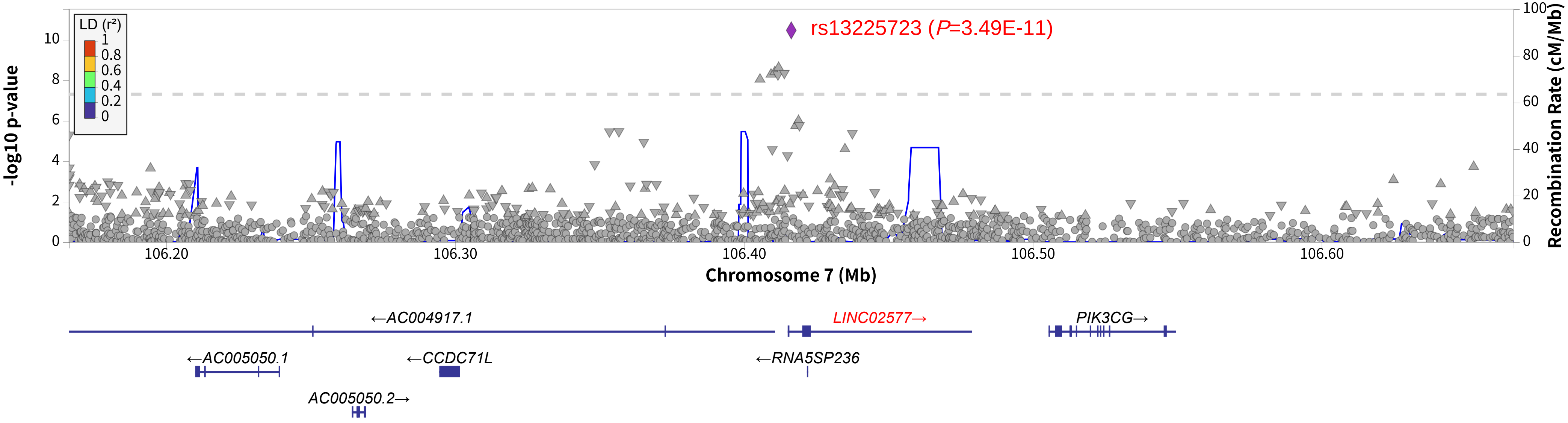


**Supplementary Fig. 8 Locuszoom plots for rs13225723 of cross-ancestry meta-analysis (stage 3, METAL) genome-wide significant association.** rs13225723 is located on *LINC02577.* The *P*-values are from fixed-effects model GWAS meta-analyses, and all association analyses were two-sided.

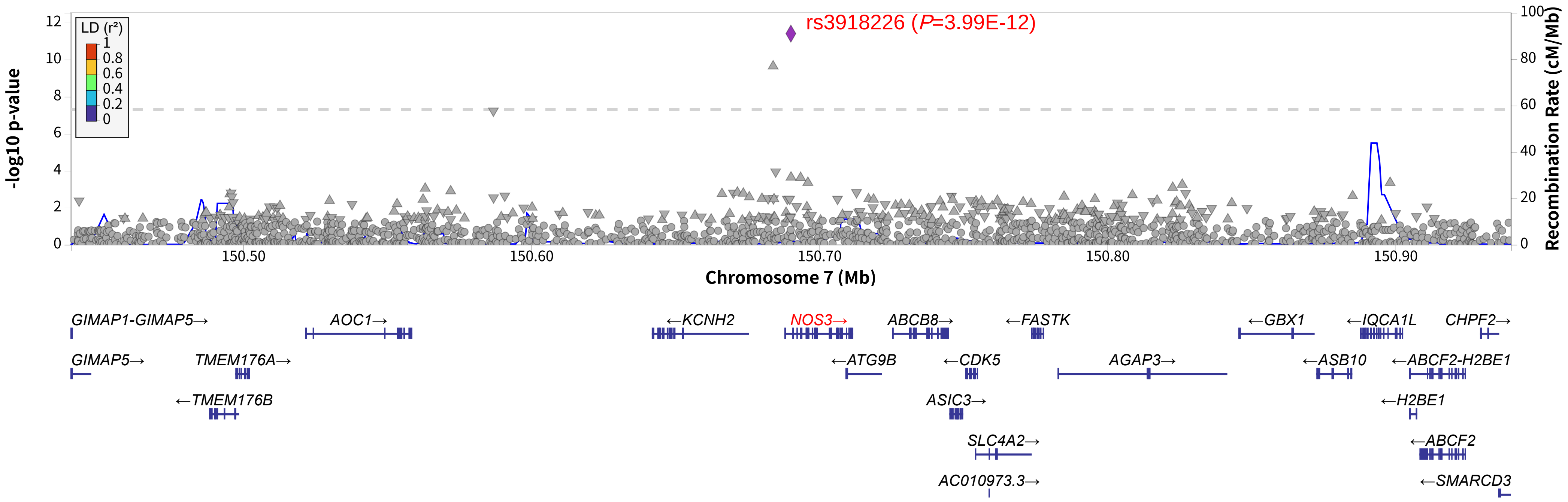


**Supplementary Fig. 9 Locuszoom plots for rs3918226 of cross-ancestry meta-analysis (stage 3, METAL) genome-wide significant association.** rs3918226 is located upstream of *NOS3*. The *P*-values are from fixed-effects model GWAS meta-analyses,and all association analyses were two-sided.

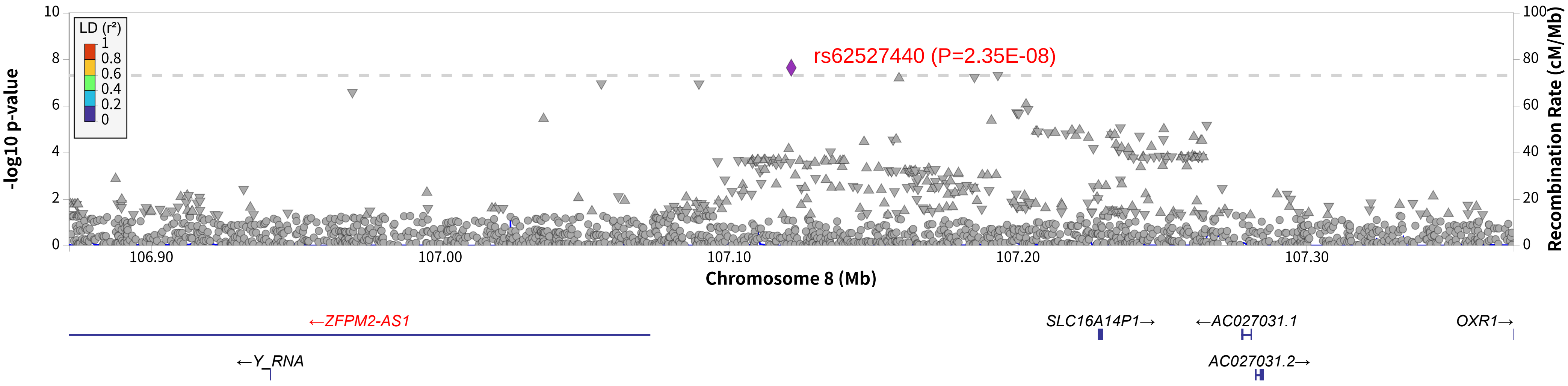


**Supplementary Fig. 10 Locuszoom plots for rs62527440 of cross-ancestry meta-analysis (stage 3, METAL) genome-wide significant association.** rs62527440is located upstream of *ZFPM2-AS1*. The *P*-values are from fixed-effects model GWAS meta-analyses,and all association analyses were two-sided.

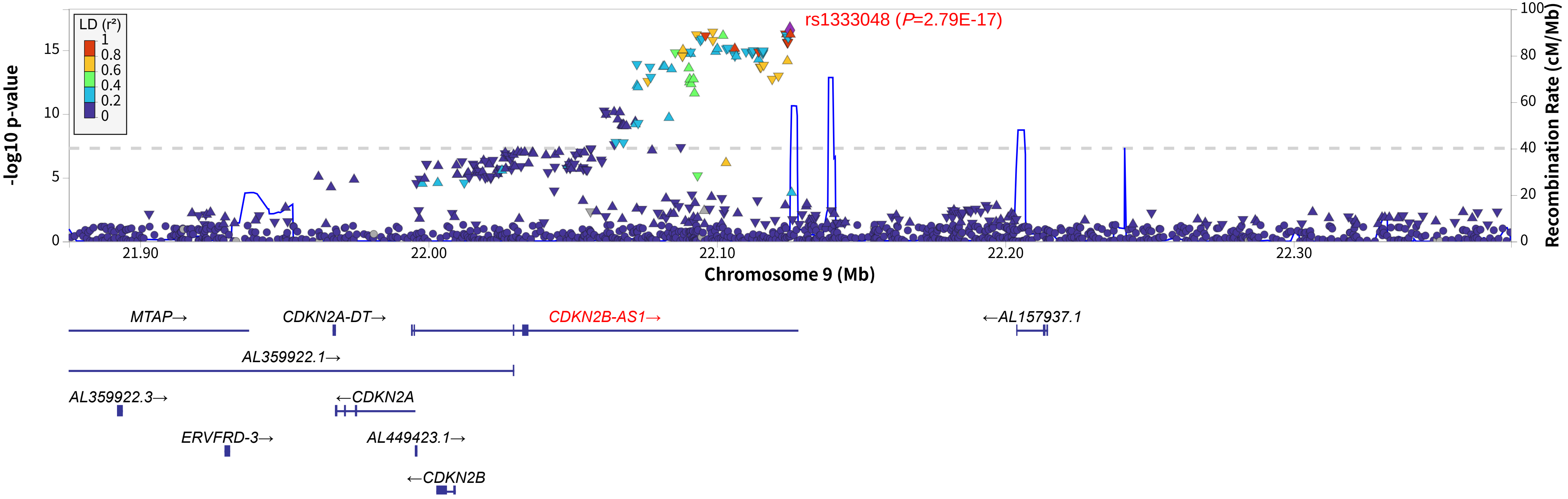


**Supplementary Fig. 11 Locuszoom plots for rs1333048 of cross-ancestry meta-analysis (stage 3, METAL) genome-wide significant association.** rs1333048is located on *CDKN2B-AS1*. The *P*-values are from fixed-effects model GWAS meta-analyses, and all association analyses were two-sided.

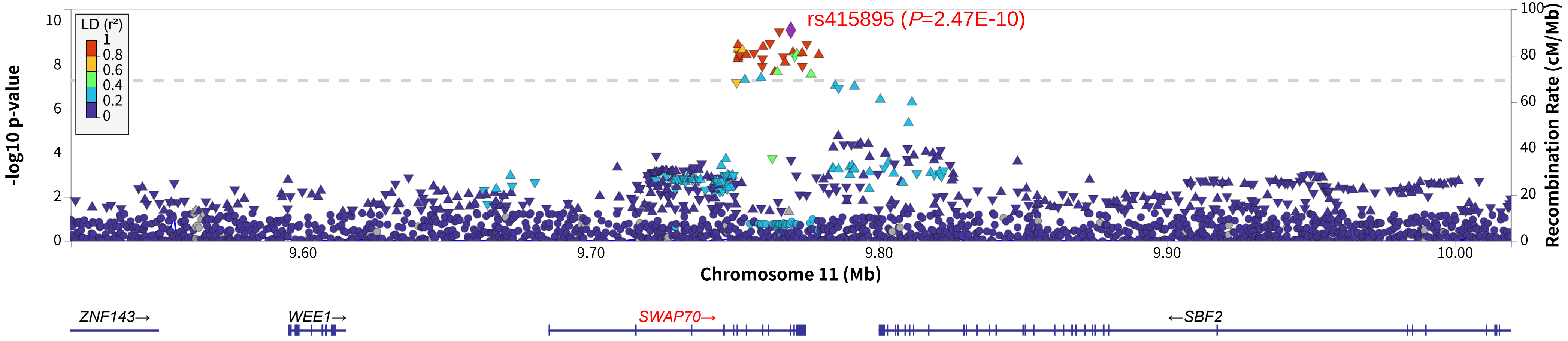


**Supplementary Fig. 12 Locuszoom plots for rs415895 of cross-ancestry meta-analysis (stage 3, METAL) genome-wide significant association.** rs415895 is located on *SWAP70*. The *P*-values are from fixed-effects model GWAS meta-analyses, and all association analyses were two-sided .

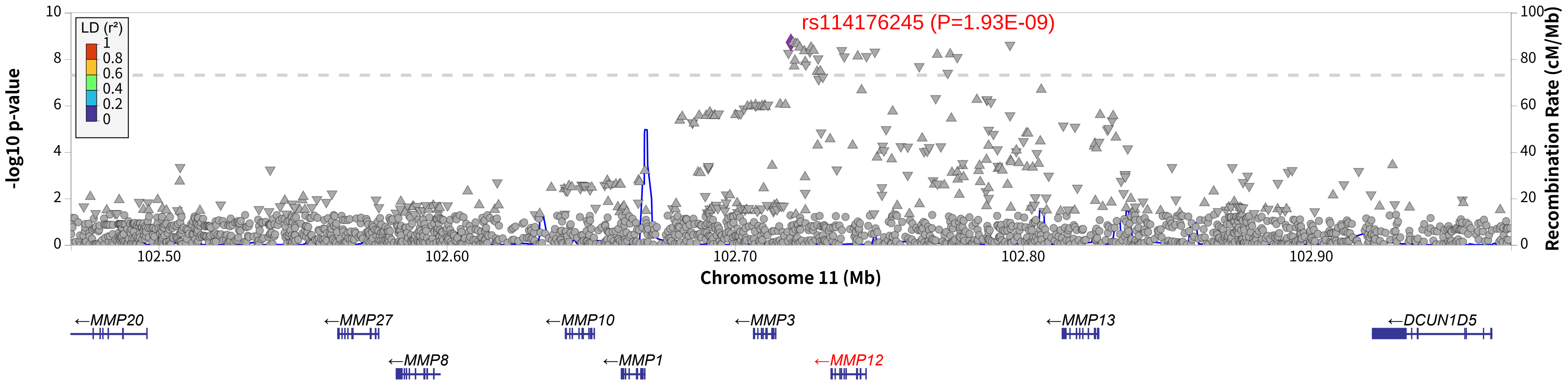


**Supplementary Fig. 13 Locuszoom plots for rs114176245 of cross-ancestry meta-analysis (stage 3, METAL) genome-wide significant association.** rs114176245 is located downstream of *MMP12*.The *P*-values are from fixed-effects model GWAS meta-analyses,and all association analyseswere two-sided.

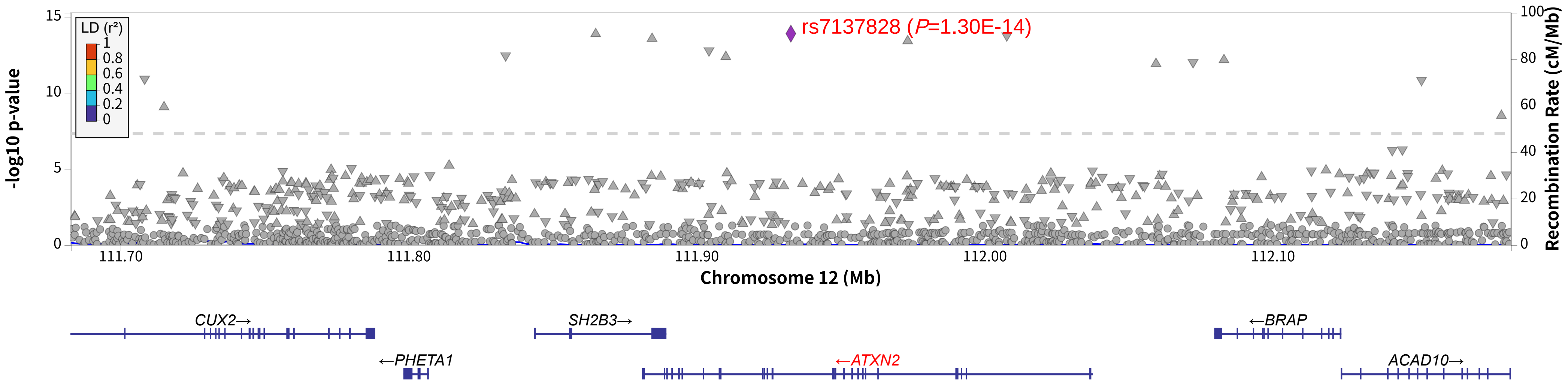


**Supplementary Fig. 14 Locuszoom plots for rs7137828 of cross-ancestry meta-analysis (stage 3, METAL) genome-wide significant association.** rs7137828 is located on *ATXN2*. The *P*-values are from fixed-effects model GWAS meta-analyses, and all association analyses were two-sided.

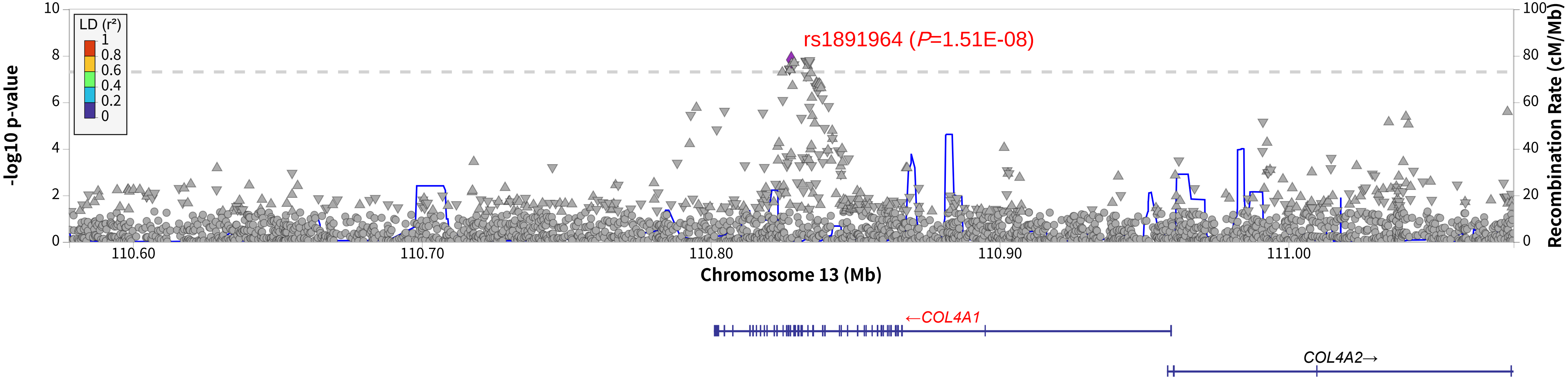


**Supplementary Fig. 15 Locuszoom plots for rs1891964 of cross-ancestry meta-analysis (stage 3, METAL) genome-wide significant association.** rs1891964 is located on *COL4A1*. The *P*-values are from fixed-effects model GWAS meta-analyses, and all association analyses were two-sided .

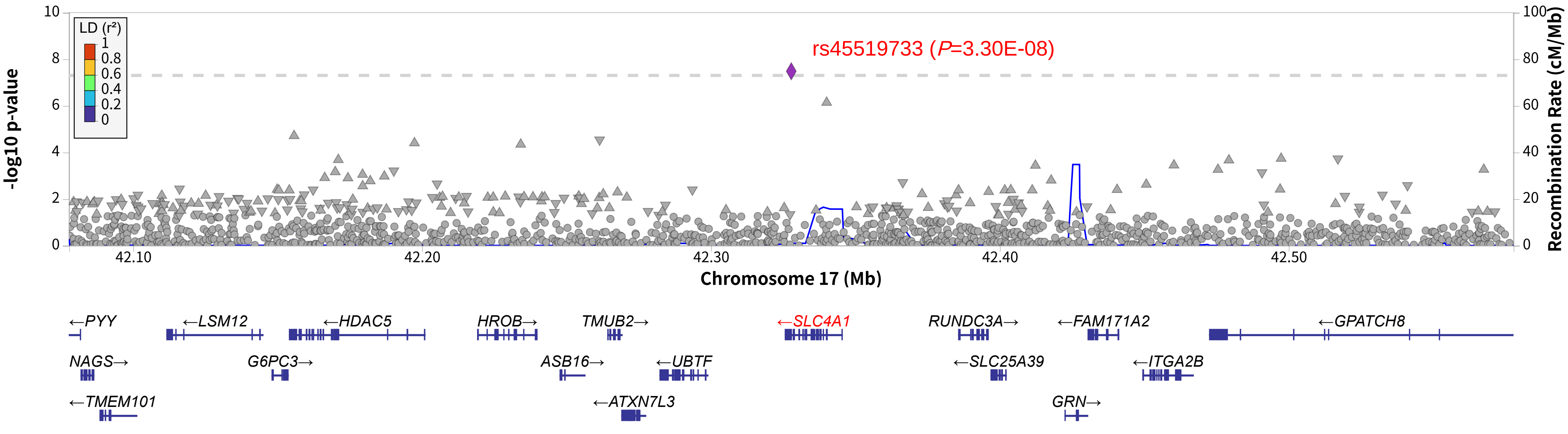


**Supplementary Fig. 16 Locuszoom plots for rs45519733 of cross-ancestry meta-analysis (stage 3, METAL) genome-wide significant association.** rs45519733 is located on *SLC4A1*. The *P*-values are from fixed-effects model GWAS meta-analyses, and all association analyses were two-sided.

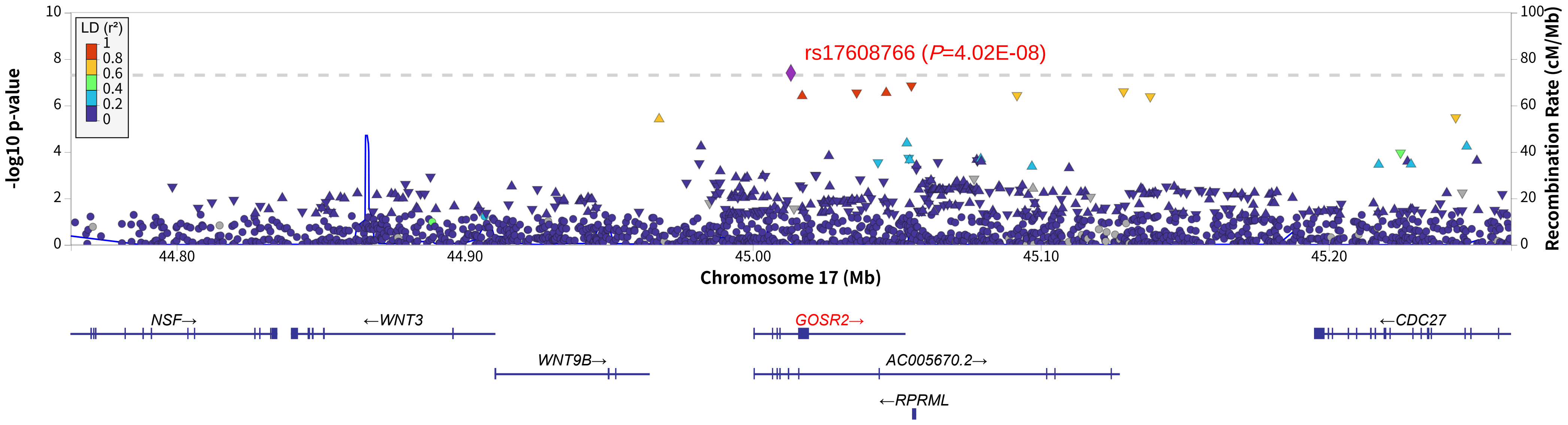


**Supplementary Fig. 17 Locuszoom plots for rs17608766 of cross-ancestry meta-analysis (stage 3, METAL) genome-wide significant association.** rs17608766is located on *GOSR2.* The *P*-values are from fixed-effects model GWAS meta-analyses, and all association analyses were two-sided.

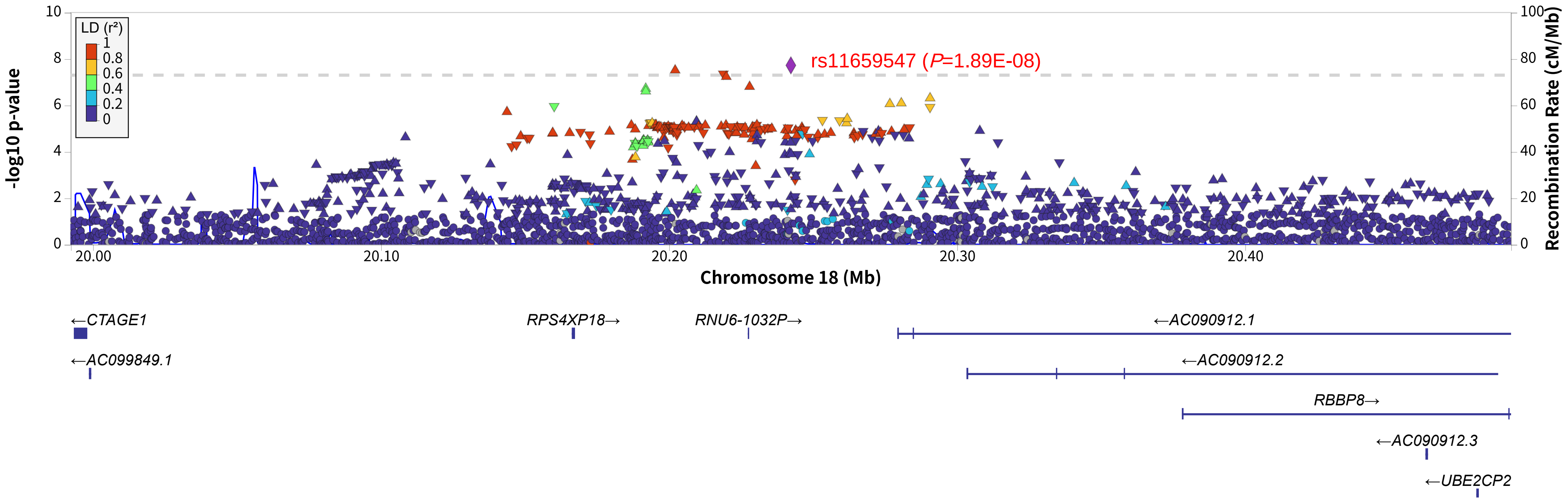


**Supplementary Fig. 18 Locuszoom plots for rs11659547of cross-ancestry meta-analysis (stage 3, METAL) genome-wide significant association.** rs11659547 is located on the upstream of *RNU6-1032P.* The *P*-values are from fixed-effects model GWAS meta-analyses, and all association analyses were
two-sided.

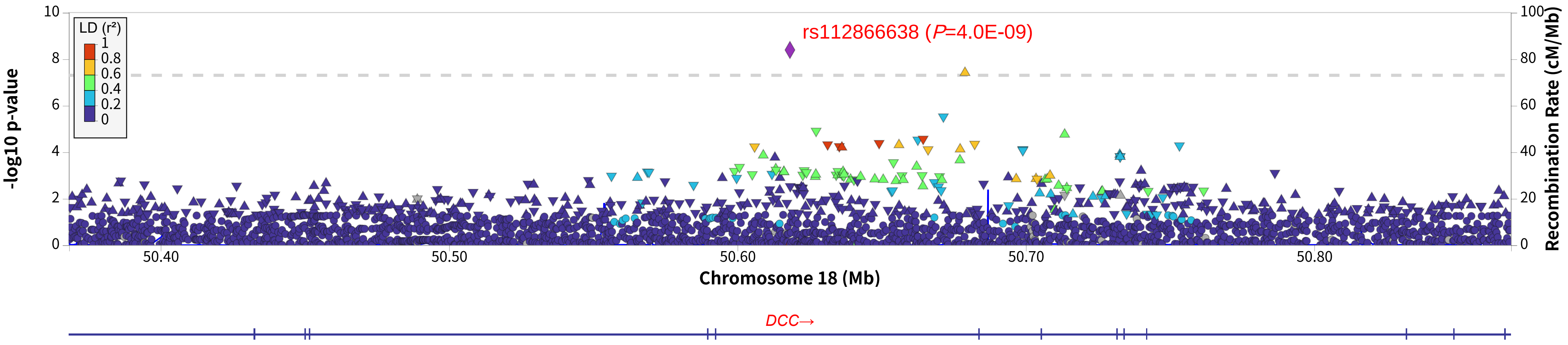


**Supplementary Fig. 19 Locuszoom plots for rs112866638 of cross-ancestry meta-analysis (stage 3, METAL) significant association.** rs4297043 is located on *DCC.* The *P*-values are from fixed-effects model GWAS meta-analyses, and all association analyses were two-sided.

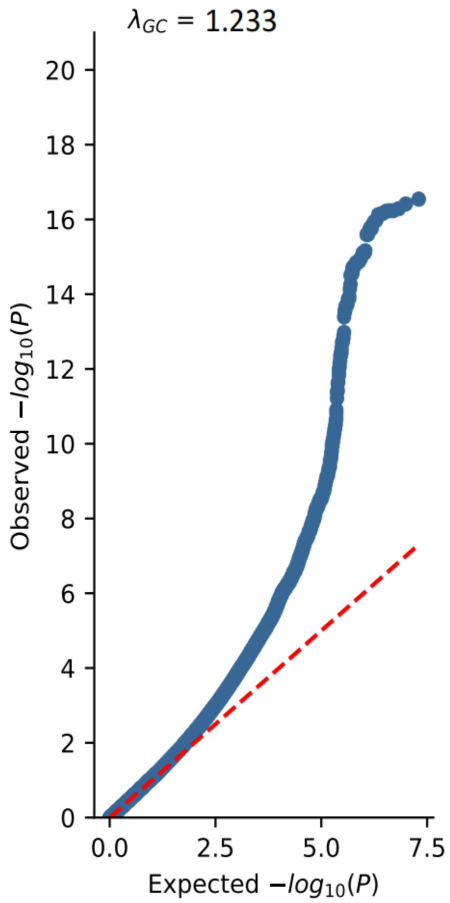


**SupplementaryFig. 20 QQ plot and LambdaGC of the cross-ancestry (stage 2, METAL) meta-analysis of TIA.** The observed *P*-values were from fixed-effects model GWAS meta-analyses,and all association analyses weretwo-sided.The red dashed line represents the predicted *P*-value, and the blue framed line shows the 95% confidence interval.

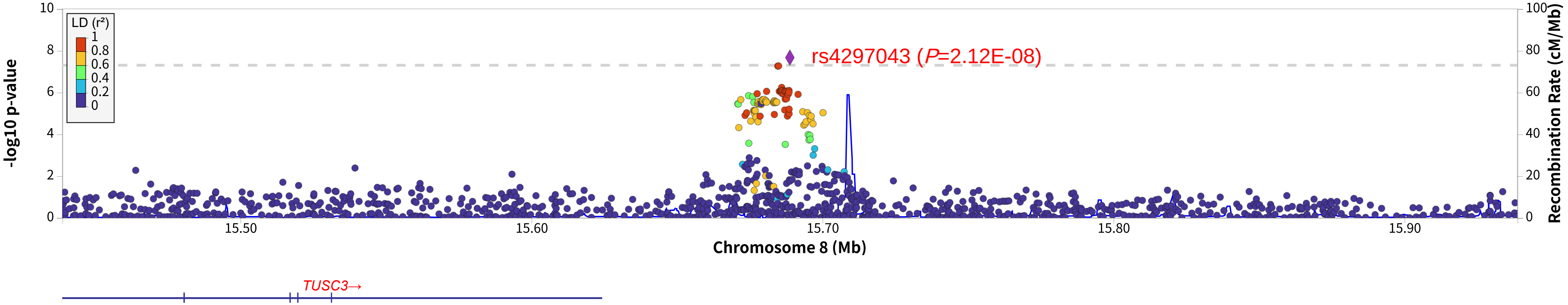


**Supplementary Fig. 21 Locuszoom plots for rs4297043 of cross-ancestry meta-analysis (stage 3, MR-MEGA) significant association.** rs4297043 is located on the downstream of *TUSC3.* The *P*-values are from fixed-effects model GWAS meta-analyses, and all association analyses were two-sided.

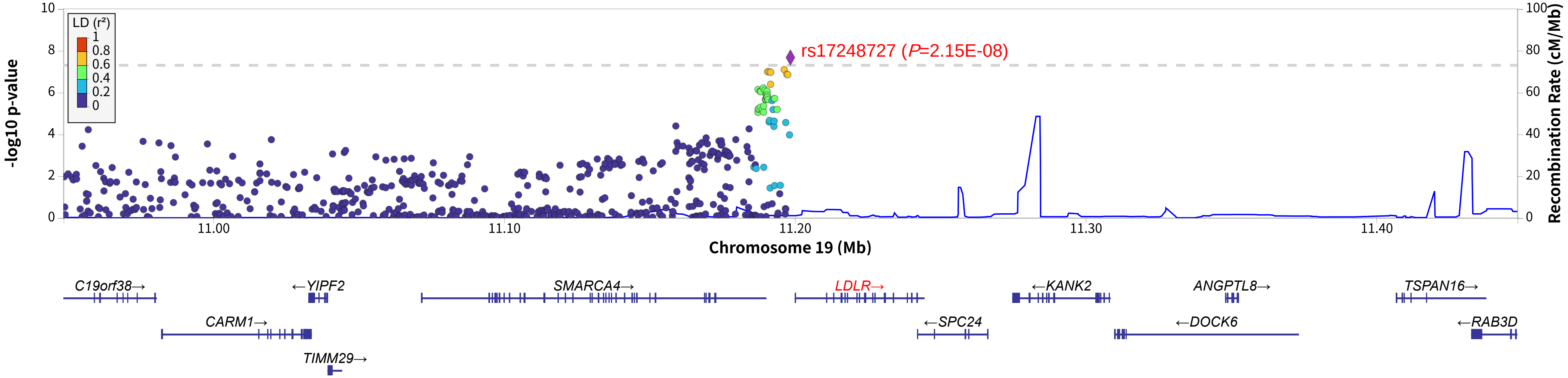


**Supplementary Fig. 22 Locuszoom plots for rs17248727 of cross-ancestry meta-analysis (stage 3, MR-MEGA) significant association.** rs17248727 is located on the upstream of *LDLR.* The *P*-values are from fixed-effects model GWAS meta-analyses, and all association analyses were two-sided.
